# EXTRACELLULAR MATRIX REMODELING IN ATOPIC DERMATITIS HARNESSES THE ONSET OF AN ASTHMATIC PHENOTYPE AND IS A POTENTIAL CONTRIBUTOR TO THE ATOPIC MARCH

**DOI:** 10.1101/2022.01.17.22269397

**Authors:** Patrick Graff, Jenny Wilzopolski, Anne Voss, Travis M. Blimkie, January Weiner, Olivia Kershaw, Preety Panwar, Tillie Hackett, Dieter Brömme, Lucie Loyal, Andreas Thiel, Dieter Beule, Robert E.W. Hancock, Achim D. Gruber, Wolfgang Bäumer, Sarah Hedtrich

## Abstract

The development of atopic dermatitis (AD) in infancy, and subsequent allergic rhinitis, food allergies, and asthma in later childhood, is known as the atopic march. The mechanism is largely unknown, yet the course of disease indicates the contribution of inter-epithelial crosstalk, through to the onset of inflammation in the skin and progression to another mucosal epithelium.

Here, we investigated if and how skin-lung epithelial crosstalk could contribute to the development of the atopic march. First, we emulated this inter-epithelial crosstalk through indirect co-culture of bioengineered atopic-like skin disease models and three-dimensional bronchial epithelial models triggering an asthma-like phenotype in the latter. A subsequent secretome analysis identified throm-bospondin-1, CD44, complement factor C3, fibronectin, and syndecan-4 as potentially relevant skin-derived mediators. As these mediators are extracellular matrix (ECM)-related proteins, we then studied the involvement of the ECM, unveiling distinct proteomic, transcriptomic, and ultrastructural differences in atopic samples. The latter indicated ECM remodeling triggering the release of the above-mentioned mediators. In addition to pro-inflammatory effects in lung tissue, the ECM mediators also exert distinct effects on CD4^+^ T cells. *In vivo* mouse data showed that exposure to these mediators over seven days dysregulated activated circadian clock genes which have been previously discussed in the context of atopic diseases and asthma development.

We hypothesize the existence of a skin-lung axis that could contribute to the atopic march driven by skin ECM remodeling.

**One Sentence Summary:** Atopic skin harbors the progression of atopic diseases to lung tissue through a skin-lung axis that contributes to the atopic march via extracellular matrix remodeling.

## Introduction

The worldwide prevalence of allergic diseases has continuously increased over the past decades posing a tremendous socioeconomic burden and significantly impacting the quality of life for atopic individuals. Atopic dermatitis (AD) is the most common inflammatory skin disease, with a point prevalence in children of up to 20% and 2-5% in adults [*1, 2*]. Importantly, AD often heralds the onset of other atopic diseases such as allergic rhinitis, eosinophilic esophagitis, food intolerance, and allergic asthma (AA); a concept known as the atopic march [*3-5*]. Although the underlying mechanisms are largely unknown, epidemiological data point towards a temporal association and causal link between these diseases [*4, 6*]. For example, the Canadian Healthy Infant Longitudinal Development (CHILD) study found that children diagnosed with AD and concomitant sensitization at an age of 1 had a 7-fold increased risk of developing AA at an age of 3-years [*7*]. Globally, the data vary considerably, but estimates suggest that 30-50% of children with moderate-to-severe AD may eventually develop AA [*4*] and patients with moderate-to-severe AD are generally at greater risk [*8*].

Further, it is clear that there is a genetic component to the atopic march, yet its contribution to disease manifestation is only partly understood. The strongest known genetic risk factor for AD are loss-of-function mutations in the filaggrin gene (*FLG*) which encodes the barrier protein filaggrin [*9*]. A lack of filaggrin result in skin barrier dysfunction facilitating the entry of allergens and thus promoting the onset of AD. Interestingly, although people with mutations in *FLG* are at increased risk to develop AA [*10*], this correlation only holds for individuals with AD [*11*], whereas people that carry *FLG* mutations, but do not suffer from AD, are not at greater risk for developing AA.

Strikingly, the skin epithelium in AD and lung epithelium in AA have highly similar disease patterns, in which they both respond to environmental allergens with a Th2-driven inflammatory response evolving around the epithelium-derived cytokine thymic stromal lymphopoietin (TSLP) as a central player [*12-16*]. TSLP is highly overexpressed in AD lesions and exerts pro-inflammatory effects by the activation and polarization of CD4^+^ and CD8^+^ T cells [*17-19*].

As the atopic march typically first manifests as AD within 6-24 months after birth and then progresses to other epithelia such as the intestine and the lungs, this points towards a role for interepithelial crosstalk. How the epithelia communicate is largely unknown. Yet, for the atopic march, the contribution of TSLP [*12, 16*], T cells [*20*], and impaired antiviral IFN production secondary to skin barrier defects [*21*] are being discussed.

It is noteworthy that studying the skin-lung crosstalk in the context of atopic diseases is hampered by the lack of suitable disease models that would allow to experimentally address the complex interplay of genetic and environmental factors, epithelial barrier defects, and immune dysregulations yielding highly heterogenous phenotypes [*6, 22*]. Although animal models of atopic diseases and the atopic march exist, their predictive value for the human situation is controversial since humans differ significantly from rodents [*4, 23, 24*]. Further, most laboratory mice do not spontaneously develop atopic diseases. In AD mouse models, the disease phenotype is induced by a broad range of methods, for example by repeated topical application of sensitizers such as oxazolone, allergens like ovalbumin, and house dust mites or by using genetically modified mice. To mimic an atopic march-like process, allergens like house dust mites or ovalbumin are administered via inhalation or s.c. injection t [*25*]. Detailed insights about the model’s capability to mimic specific aspects of AD are described elsewhere [*26, 27*]. To our knowledge, none of the existing mouse models of AD emulate the whole process of the atopic march from skin to other organs. Lastly, there is poor homology between the transcriptomic profile of murine AD models and patients with AD [*28*].

To understand the potential contribution of skin-lung epithelial crosstalk in the atopic march, we employed a bioengineered human atopic-like skin disease model, that emulates characteristics of AD, and investigated its effects on 3D bronchial epithelial models. We identified five mediators (thrombospondin-1, CD44, complement factor C3, fibronectin, and syndecan-4) as potentially relevant. The majority of these factors were extracellular matrix (ECM)-related proteins that prompted us to further investigate the contribution of dermal ECM remodeling to lung inflammation. Hence, we conducted multi-step proteomic and transcriptomic analyses showing significantly different expression profiles on gene and ECM level between normal and AD patient-derived fibroblasts. Our investigations were complemented by immunohistological analysis, ELISA, RT qPCR, and western blot data of AD patient-derived samples, primary CD4^+^ and CD8^+^ T cells, and *in vivo* mouse studies pointing towards a skin-lung crosstalk mediated by skin ECM-derived mediators. Interestingly, sub-chronic exposure of mice to these ECM-derived mediators significantly dysregulated circadian clock genes which are known players in epithelial inflammation and targets-of-interest for asthma therapy.

## Results

### Atopic-like skin models triggered an asthma-like phenotype in bronchial epithelial models

To assess the effects of a human atopic-like skin disease model on a healthy bronchial epithelium, we cultivated the latter in cell culture medium that was previously collected from atopic skin models **(Fig. 1A)**. This indirect co-culture triggered an inflammatory phenotype in the bronchial epithelium, that was characterized by hyperproliferation, increased mucus production, upregulation of alpha smooth muscle actin (*α*-SMA), MUC5AC, TSLP, squamous cell carcinoma antigen 1 (SCCA-1), the tight junction (TJ) protein zonula occludens 1 (ZO-1), and uteroglobin (UG) in the epithelial layer and tenascin C in the basal layer **(Fig. 1C)**. Complementary western blot (WB) and qRT PCR data are shown in **Fig. 2A-C**. Concordantly, increased mucus production and *α*-SMA expression were also confirmed in lung tissue sections from asthmatics, and bronchial epithelial models generated from asthma patient cells **(Fig. 1B)**.

**Figure 1.**
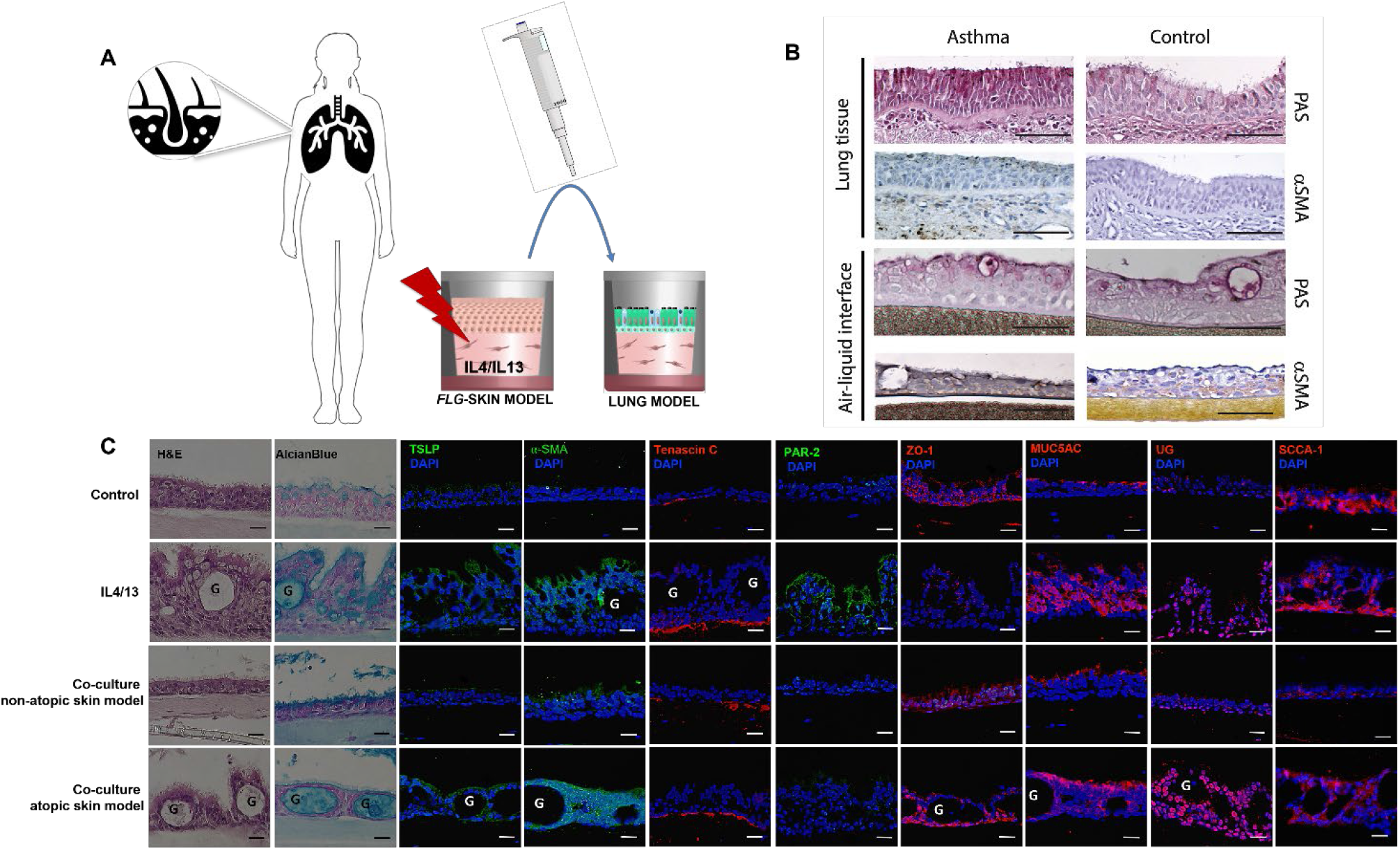
A) Schematic drawing of the indirect skin-lung co-culture model: Cell culture medium collected from healthy (=non-atopic) or diseased (=atopic, *FLG-*_IL-4/IL-13_) skin models was collected, mixed with 50% fresh medium and added to bronchial epithelial models every other day for 6 days. **B) Representative periodic acid-Schiff (PAS)-, and *α*-SMA-IHC staining** of healthy and asthmatic human lung tissue sections and patient-derived air-liquid interface bronchial epithelial models. Scale bar: 100 μm. **C) Representative H&E-, mucus alcian blue- and immunofluorescence-staining** against TSLP, alpha smooth muscle actin (*α*-SMA), tenascin C, PAR-2, ZO-1, MUC5AC, uteroglobin (UG), and SCCA-1 of bronchial epithelial models after indirect co-culture with non-atopic and atopic-like skin models as well as IL-4/IL-13 over 6 days. G = goblet cells. Scale bar: 25μm. Exposure times: blue channel 1/15s, red channel 1/3s, green channel 1/3s. Scale bar = 25μm.

**Figure 2.**
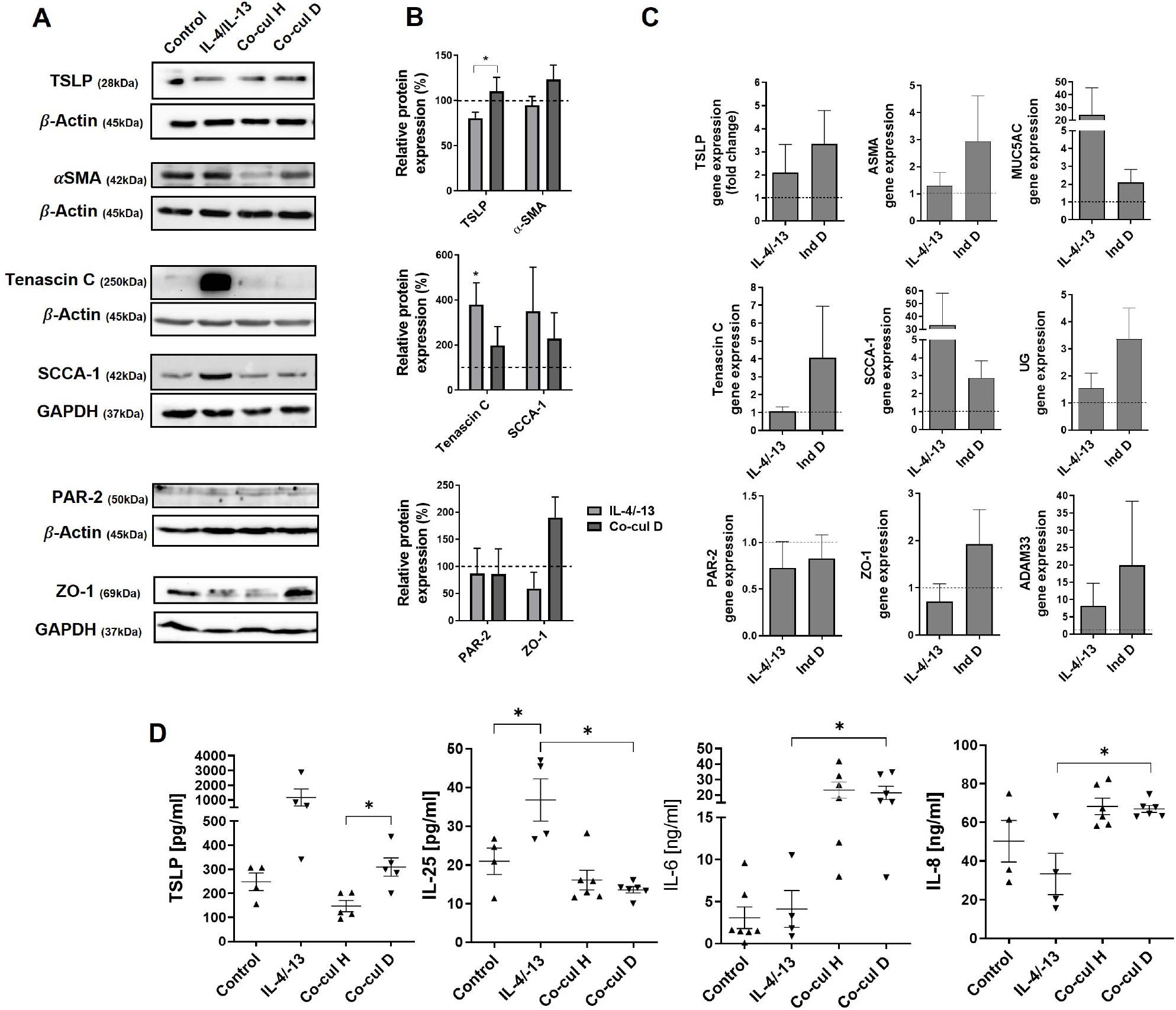
Protein, relative gene expression, and cytokine analysis of bronchial epithelial models after indirect co-culture with non-atopic (Co-cul H) and atopic-like (Co-cul D) skin models as well as IL-4/IL-13. **A)** Representative western blot bands of TSLP, α-SMA, tenascin C, SCCA-1, PAR-2, and ZO1 in bronchial epithelial models were semi-quantified by densitometry analysis **(B)** using Image J (Fiji Version 1.0), mean ± SEM, n = 3-5. **C)** Corresponding relative gene expression data depicted as fold change of *TSLP, ASMA, MUC5AC*, tenascin C, *SCCA-1*, uteroglobin (*UG*), *PAR-2, ZO-1*, and *ADAM33* to respective untreated control (dotted line) in bronchial epithelial models as determined by qRT-PCR. *GAPDH* served as housekeeping gene. Mean ± SEM, n=3. **D)** TSLP, IL-25, IL-6, and IL-8 cytokine levels in cell culture media of bronchial epithelial models after 6-days indirect co-culture with non-atopic and atopic-like skin models or treatment with IL-4/IL-13 as determined by sandwich ELISA. n=5-6 independent donors. *p ≤ 0.05, paired t-test. Co-cul H: co-culture with non-atopic skin models; Co-cul D: co-culture with atopic-like skin models.

An IL-4/IL-13 control group was included for reference. As expected, the addition of IL-4/IL-13 to the culture medium of bronchial epithelial models also caused an inflammatory phenotype, although the effects were distinct from the effects of the skin model-derived medium. In fact, IL-4/IL-13 yielded a more pronounced upregulation of tenascin C and SCCA-1, yet had no or little effect on ZO-1, TSLP, UG, and *α*-SMA expression.

We then quantified atopy-relevant cytokines in the culture medium of the bronchial epithelial models after indirect co-cultivation and found significantly increased TSLP secretion in line with the WB data (**Fig. 2B**), yet no increase in IL-25, IL-6, and IL-8 levels, and no IL-33 compared to the co-culture with non-atopic skin models. In contrast, IL-4/IL-13 supplementation triggered significant IL-25 release (**Fig. 2D)**.

### Skin Matrisome-Associated Mediators Triggered Inflammation in Bronchial Epithelial Models

Next, we ran a secretome analysis of the culture medium collected from the atopic-like skin models to identify secreted factors that may have contributed to the inflammatory phenotype in the bronchial epithelial models (**Fig. 3A)**. Aiming for a targeted search, we first ran an SDS gel to assess differences in the protein band profiles of culture medium from normal and atopic-like skin models. The gel electrophoresis revealed two bands of interest, at ∼150 kDa and ≥250 kDa, respectively, that were either absent in the medium of non-atopic skin models or stronger in the culture medium of atopic-like models **(Fig. 3B)**.

**Figure 3.**
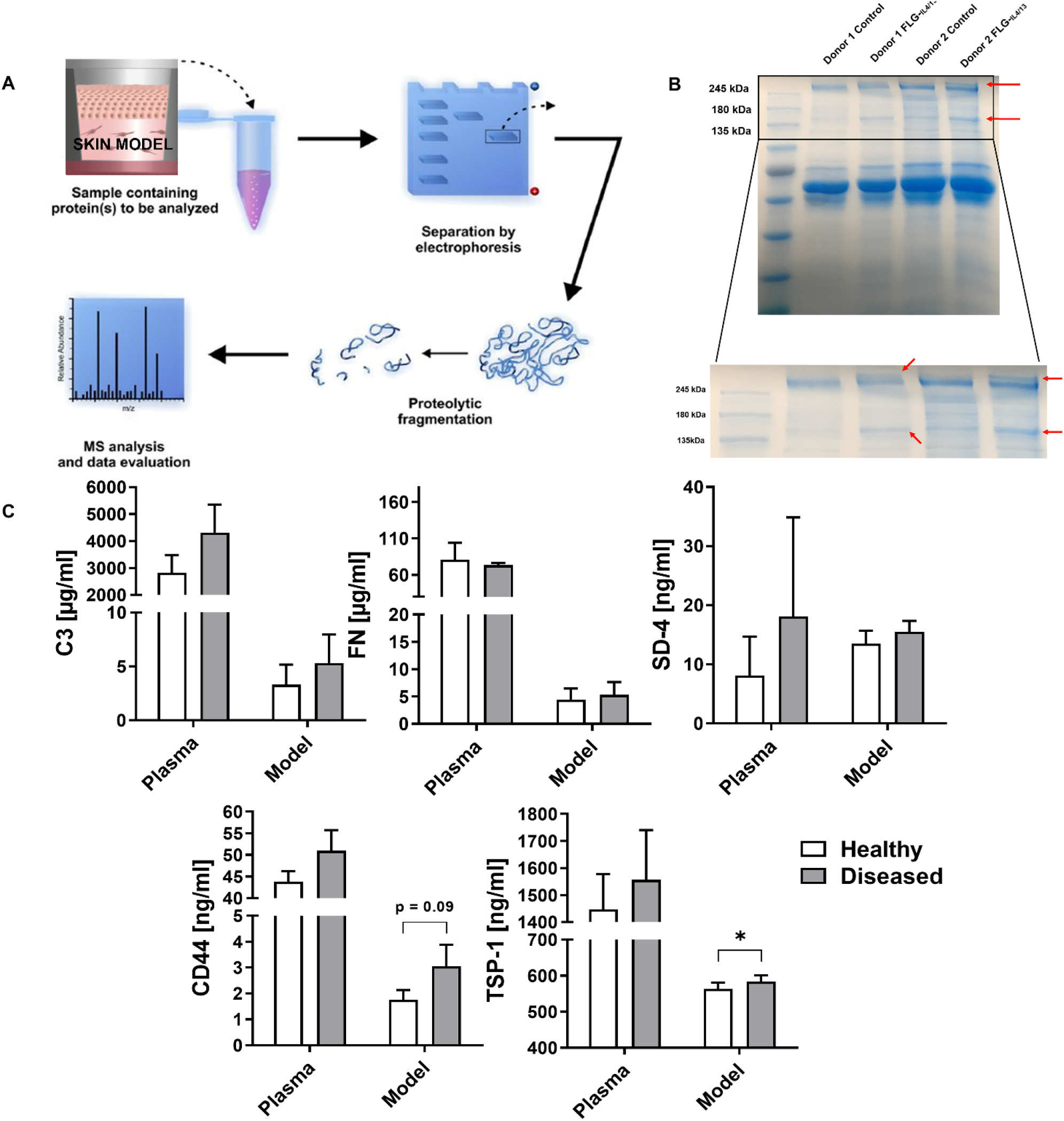
Secretome analysis of skin models. **A)** Schematic drawing of the secretome analysis approach. Supernatants collected from non-atopic and atopic-like (FLG-_IL-4/IL-13_) skin models were subjected to SDS gel electrophoresis and stained with coomassie blue. Subsequently, bands of interest were cut out and analyzed by mass spectrometry. Adapted and reprinted from [*29*] under the terms and conditions of the Creative Commons Attribution license (Attribution 3.0 Unported, CC BY 3.0). **B)** Representative SDS gel showing differences (red arrows) between non-atopic and atopic-like (FLG-_IL-4/IL-13_) skin models in bands with a size of ∼150 kDa and ∼250 kDa. **C)** C3, fibronectin (FN), syndecan-4 (SD-4), CD44, and thrombospondin-1 (TSP-1) levels in cell culture media of non-atopic and atopic-like (FLG-_IL-4/IL-13_) skin models and blood plasma from 5-year old children diagnosed with atopic dermatitis and eosinophilic esophagitis as second manifestation of the atopic march. Plasma samples from non-atopic children served for control. Mean ± SEM. n=5 independent donors for blood samples, n=7 independent donors for skin models. * p ≤ 0.05, paired T-test.

Next, we cut out those bands and subjected them to proteomic analysis which provided us with lists of 20-50 proteins per biological repeat (**Data Set S1**). In addition to classic ECM-related proteins such as different collagen subtypes of collagen, keratin, and laminin, we classified complement factor C3 (C3), fibronectin (FN), syndecan-4 (SD-4), cluster of differentiation 44 (CD44), and thrombos-pondin-1 (TSP-1) as potentially relevant. It should be noted that both CD44 and SD-4 are proteoglycans and therefore heterogeneously glycosylated which explains their appearance in these high MW fractions. Interestingly, all these mediators have been linked to the skin extracellular matrix (ECM) which prompted us to run more detailed analysis on the involvement of skin ECM.

We then quantified the five mediators in the cell culture medium of our skin models and in blood plasma samples from children (median age 5 years) that have been diagnosed with AD and eosinophilic esophagitis (EoE). The latter is considered a late stage complication of the atopic march [*5*]. Plasma samples from age-matched children without atopic diseases served as control. In the culture medium of the atopic-like skin models, we found significantly increased levels of TSP-1 relative to non-atopic disease controls. For CD44 (p = 0.09), C3 (p = 0.22), SD-4 (p = 0.21), and FN (p =0.47) a similar trend was observed (**Fig. 3C**). A clear tendency towards increased plasma levels for all mediators, except FN, was also detected in the plasma samples from atopic children, yet without statistical significance **(Fig. 3C**). The latter is likely due to the low sample size and the retrospective analysis of the samples that were retrieved from a biobank. The selection criteria for our study were children diagnosed with AD and another atopic disease (= atopic samples) that manifests as part of the atopic march versus blood samples from non-atopic children. Yet, no further clinically relevant information such as disease severity, age of onset etc. had been available.

To examine the effects of each mediator on lung tissue, we then directly added C3, CD44, TSP-1, SD-4, and FN to the culture medium of normal bronchial epithelial models. Similar to the culture medium of the atopic-like skin models (**Fig. 1**), all mediators triggered a hyper-proliferative phenotype with hyperplasia of mucus-producing goblet cells, and CD44, TSP-1, SD-4, and FN an increase in TSLP, α-SMA, tenascin C, and ZO-1 expression (**Fig. 4A, S1**). Again, these findings were confirmed by WB analysis and RT qPCR (**Fig. 4B, Fig. S2**). Subsequently, we also quantified the levels of IL-6, IL-8, IL-25, and TSLP in the culture medium of the bronchial epithelial models, yet no significant effects were detected (**Fig. S2**) indicating that the culture medium of atopic skin models contain other, so far unidentified mediators that contribute to the pro-inflammatory effects observed in **Fig. 1**.

**Figure 4.**
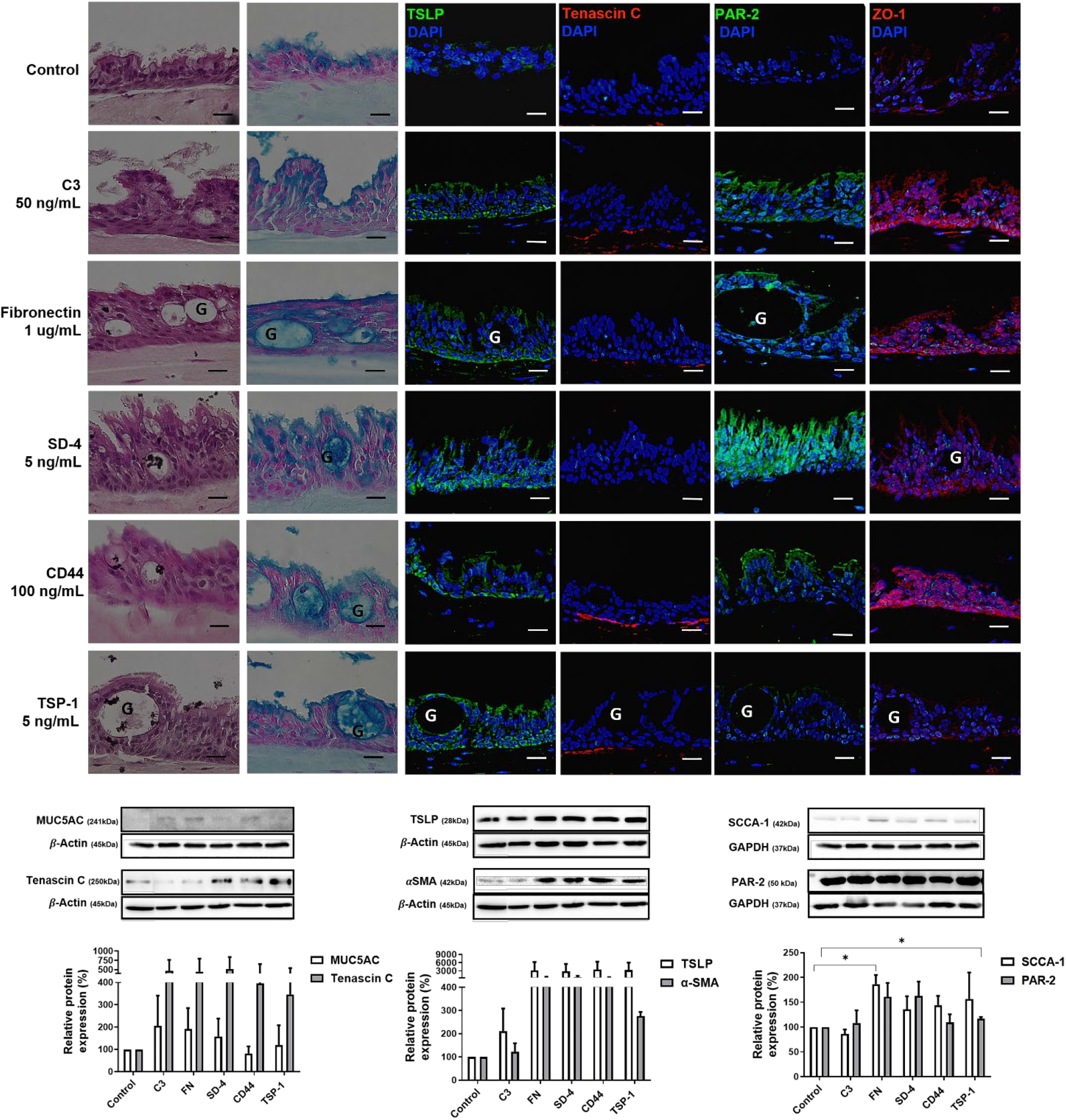
A) Representative histological, alcian blue, and immunofluorescence staining against TSLP, tenascin C, PAR-2, and ZO-1 in bronchial epithelial models are shown after treatment with 50 ng/ml complement factor C3 (C3), 1 ug/ml fibronectin (FN), 5 ng/ml syndecan-4 (SD-4), 100 ng/ml soluble cluster of differentiation 44 (CD44), or 5 ng/ml thrombospondin-1 (TSP-1) over 6 days. G = goblet cell. Scale bar = 25μm. Exposure times: blue channel 1/15s, red channel 1/3s, green channel 1/3s. **B) Corresponding western blot and relative protein expression data** for MUC5AC, tenascin C, TSLP, α-SMA, SCCA-1, and PAR-2 after 6-days treatment with 50 ng/ml C3, 1 μg/ml FN, 5 ng/ml SD-4, 100 ng/ml soluble CD44, or 5 ng/ml TSP-1 as derived from using Image J (Fiji 1.0). β -actin or GAPDH served as housekeeper. Mean ± SEM. n = 3 independent donors. * p ≤ 0.05, paired T-test.

### AD patient-derived fibroblasts produced disorganized ECM that differed significantly from normal ECM

As FN, SD-4, CD44, and TSP-1 are extracellular matrix (ECM)-related proteins, we next analysed the properties of skin ECM in normal and diseased states in more detail. We therefore stimulated ECM production in fibroblasts derived from non-atopic donors or AD patients. After 3 weeks, we obtained a three-dimensional opaque, mechanically stable ECM which was then ultrastructurally examined by scanning electron microscopy (SEM) **(Fig. 5A)**. The dermis from excised human skin served for reference. SEM analysis showed well organized collagen bundles in both, ECM generated from non-atopic individuals and that from native human skin. In contrast, ECM produced by AD fibroblasts was characterized by disorganized and less dense collagen bundles (**Fig. 5A**). Also, collagen and elastin content were significantly lower in atopic ECM samples compared to non-atopic and native skin (**Fig. 5B**) [*30*].

**Figure 5.**
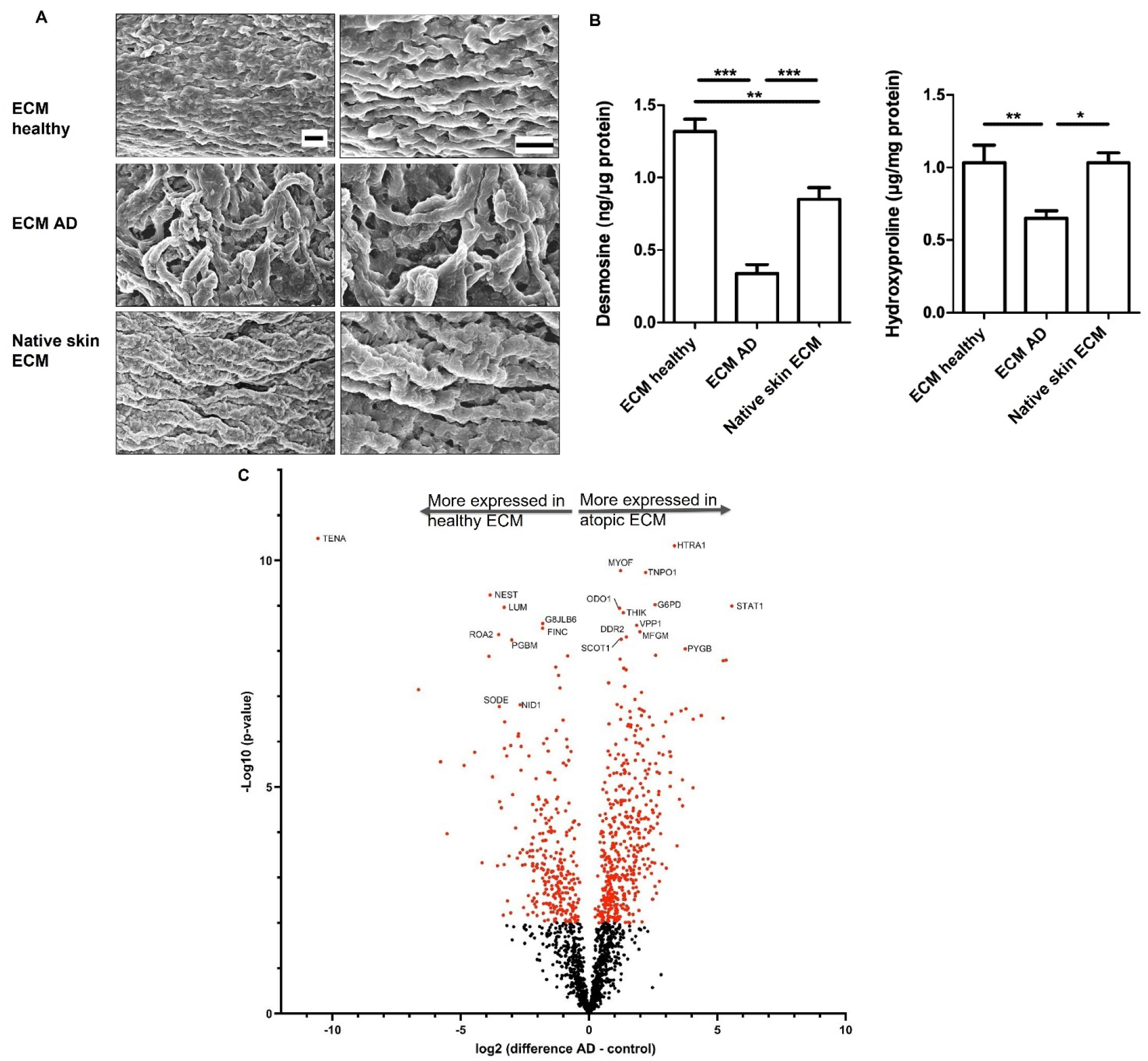
Scanning Electron Microscopy (SEM) of Skin ECM. **A)** SEM was utilized to structurally visualize ECM produced by healthy (ECM healthy), AD patient-derived fibroblasts (ECM AD), and native human skin. Scale bar= 5μm. **B) Quantification of hydroxyproline and desmosine** as indicator for collagen and elastin content in ECM. One-way ANOVA was performed, *p ≤ 0.05, **p≤ 0.01, ***p≤0.001. **C) Proteomics analysis of ECM** produced by healthy or AD patient-derived fibro-blasts. 21 proteins with the highest difference between normal and atopic ECM or high AD relevance are highlighted. Higher expression of proteins in healthy ECM is expressed by a left shift and of those in atopic ECM by a right shift. Red dots indicate *p≤ 0.01. Mean ± SEM. n=3-6 independent experiments.

For more detailed information, we next ran a semi-quantitative proteomics analysis to compare the composition and abundance of ECM proteins between non-atopic and atopic ECM samples revealing a large number of differently expressed proteins (**Data Set S2, Fig. 5C**). 21 proteins that either showed the highest differences or have a direct connection to atopic diseases are depicted in **Fig. 5** and **Table S3**. Classic ECM proteins such as tenascin, fibronectin, nestin, lumican, and the extra-cellular superoxide dismutase (SODE) were significantly more abundant in non-atopic ECM samples. In contrast, in atopic ECM samples proteins contributing to ECM remodeling and inflammation were significantly more expressed such as the serine protease HTRA1, discoidin domain-containing receptor 2, lactadherin as well as the fibroblast-related protein signal transducer and activator of transcription 1-alpha/beta (STAT1).

### AD patient-derived fibroblasts are characterized by distinct transcriptomic changes related to ECM remodeling

Based on the significant differences in protein abundancies, we next performed a transcriptomic analysis comparing atopic and non-atopic fibroblasts. This approach identified 1,593 significantly differentially expressed (DE) genes (**Data Set S3)**, revealing the considerable changes in transcription between the two groups. Pathway enrichment was used for functional analysis of these DE genes, producing 35 Reactome pathways and 20 KEGG pathways (**Data Set S4**). **Figure 6A** includes 13 key pathways and the genes within each pathway which are significantly dysregulated. These pathways have high relevance for AD, most of which demonstrate a direct connection to ECM remodeling, ECM-receptor interactions, cytokine signaling, and VEGF signaling pathways.

**Figure 6.**
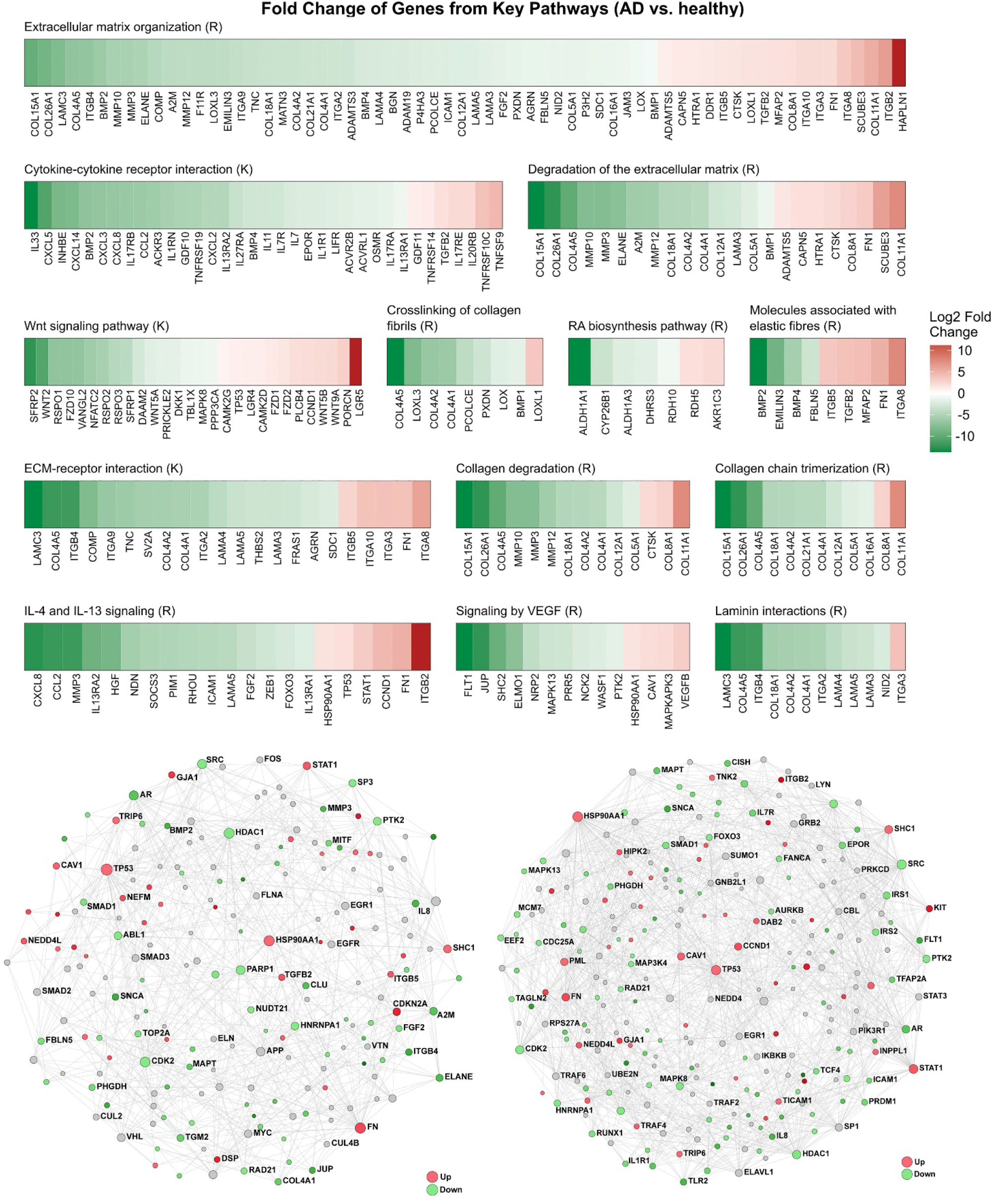
Functional analysis of transcriptomic data identifies key genes, and ECM and signaling pathways involved in AD response. **A)** Fold changes of DE genes in key Reactome and KEGG pathways identified from pathway enrichment with Sigora. An “R” or “K” denotes the source of the pathway, Reactome or KEGG, respectively. **B)** Minimally-connected first subnetworks created by extracting nodes belonging to the indicated pathway from the initial PPI network built with the 1,593 DE genes; grey nodes directly interconnect differentially expressed nodes. Key nodes (termed hubs shown by larger circles) with biological implications for AD, that were present in one or both networks included fibronectin *FN1*, heat shock protein *HSP90AA1*, transcription factor *STAT1*, integrin β5 *ITGB5*, and signal transduction adapter *SHC1*.

These DE genes were then uploaded to NetworkAnalyst to construct a minimum order protein-protein interaction (PPI) network. The network produced from this analysis was extremely dense and highly interconnected, containing more than 2,000 nodes (proteins), demonstrating the strong biological response captured in this transcriptomic data. From this large initial network, we extracted subnetworks which represent individual biological pathways identified via Reactome pathway enrichment within NetworkAnalyst. **Figure 6B** includes two of these subnetworks; “ECM Organization” and “Cytokine Signaling”, both of which show significant dysregulation and a high level of connectivity among seed nodes (DE genes). This network analysis identified genes including *FN* (fibronectin 1, a key component of the ECM), *ITGB5* (Integrin Beta-5, involved in cell adhesion), *TP53* (tumor protein 53; regulating circadian clock genes), and *STAT1* (Signal Transducer and Activator of Transcription 1, mediator of cellular response to interferons and cytokines), amongst many others, as key hubs (representing nodes that are highly interconnected within the network and thus thought to serve as key trafficking molecules that accept and disperse signals) within these highly interconnected PPI networks. Genes involved in “TLR4 Signaling,” such as *TRAF4* (TNF Receptor Associated Factor 4, signal transducer that plays a role in the activation of NF-κβ), *TLR2* (Toll-Like Receptor 2), and *IL8* were also identified as being significantly dysregulated (Figure S3).

### Skin matrisome-associated factors exert downstream effects on CD4^+^ T cell subtypes and modulate the expression of circadian clock genes

As imbalances of T helper cell subsets contribute to atopic diseases, we next interrogated the impact of the skin-matrisome associated mediators on T cells *in vitro* and *in vivo*. T cells play a central role in the pathogenesis of atopic diseases and extensive T cell activation and migration driven by leaky epithelial barriers contribute to the atopic march [*31*]. First, we assessed the effects of SD-4, CD44, and TSP-1 in male and female Balb/c mice to gain more insights into immediate effects *in vivo*. Therefore, we continuously administered SD-4, TSP-1, or CD44 over 7 days using portable pumps that were subcutaneously implanted onto the backs of the mice. The concentrations were chosen based on previously published plasma concentrations and pharmacokinetic data yielding plasma levels that were 50% above normal murine levels to avoid highly acute phenotypes. FN and C3 were excluded from this experiment due to solubility and practicability issues. After 7 days, the mice were euthanized and lung (**Fig. S4**) and spleen were removed for further analysis.

The spleens of all treated animals showed a trend towards a weight increase (PBS-treated control: 144.5 ± 26.4 mg, SD-4: 158.4 ± 9.9 mg; TSP-1: 167.3 ± 21.1 mg), including a significant increase after CD44 treatment (183.2 ± 31.2 mg). Analyzing the effects of SD-4, CD44, and TSP-1 on murine CD4^+^ T cells by assessing genetic markers for Th1 (TBX21), Th2 (GATA3), Th17 (RORC), and Th22 (IL-22) did not provide conclusive results (**Fig. S5**).

We then investigated if the five mediators exert effects on primary human naive CD4^+^ and CD8^+^ T cells on gene and protein level. We first determined the expression of subset-specific T cell markers (Th1/Tc1 (TBX21), Th2/Tc2 (GATA3), Th17/Tc17 (RORC), and Th22/Tc22 (AHR)) at the gene expression level. While no significant increase in the mRNA levels of any of these markers were found in CD8^+^ T cells, all mediators polarized and/or expanded CD4^+^ T cells. Complement factor C3, FN, and SD-4 triggered a distinct increase of TBX21, RORC, and AHR mRNA levels (**Fig. 7A**). Hardly any effect was observed on GATA3 expression. mRNA levels of naive and once (CD3/28) stimulated CD4^+^ and CD8^+^ T cells are shown in **Fig. S6**.

**Figure 7.**
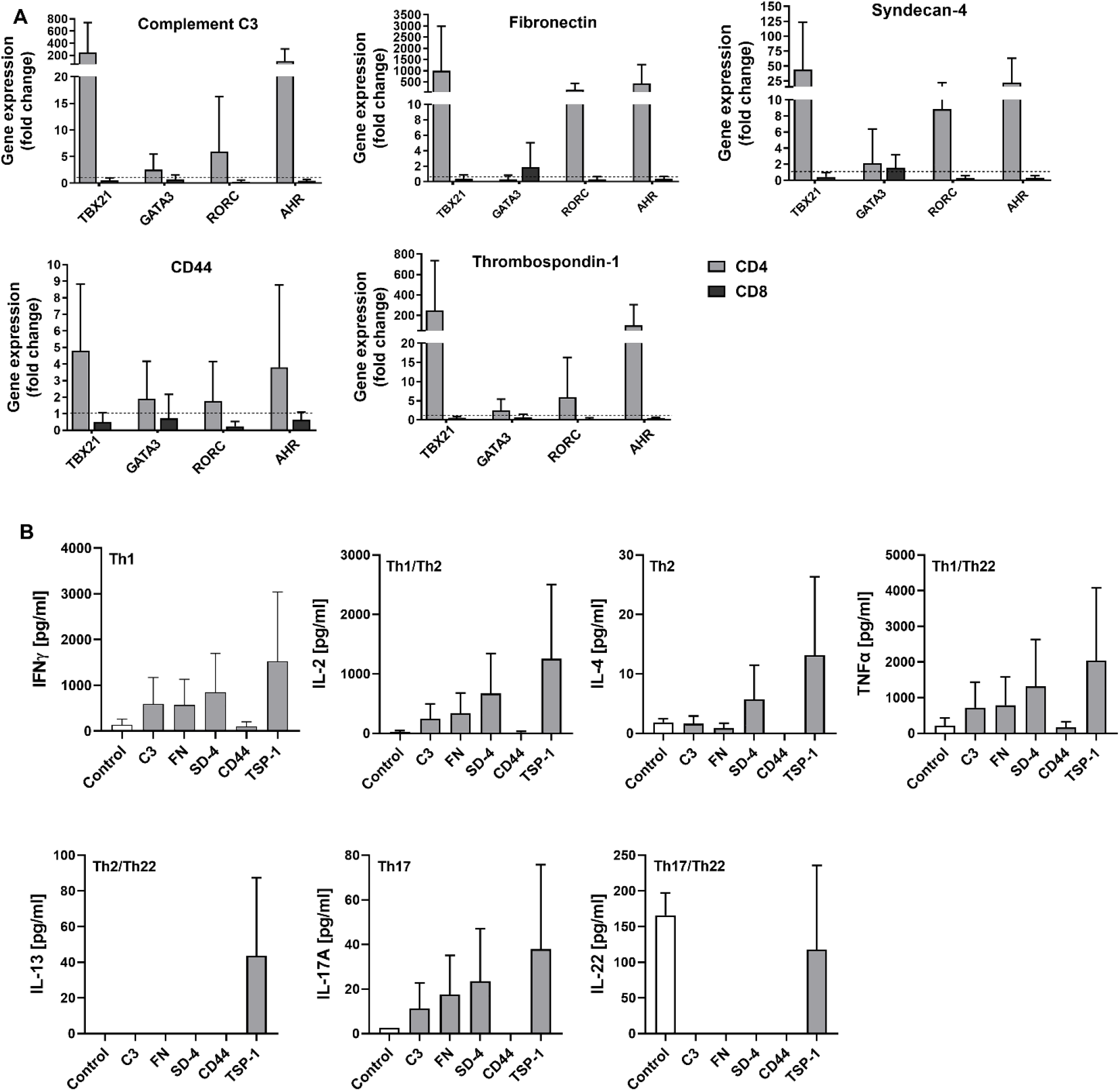
Gene expression analysis and multiplex ELISA assay of human naive CD4^+^ and CD8^+^ T cells after incubation with C3, fibronectin (FN), syndecan-4 (SD-4), CD44, and thrombos-pondin-1 (TSP-1). **A)** After isolation of naive CD4^+^ and CD8^+^ T cells from human buffy coats, CD3/CD28-activated T cells were treated with 50 ng/ml complement factor C3, 1μg/ml FN, 5 ng/ml SD-4, 100 ng/ml CD-44, and 5 ng/ml TSP-1 for 10 days. Normalized gene expression of TBX21 (Th1/Tc1), GATA3 (Th2/Tc2), RORC (Th17/Tc17), and AHR (Th22/Tc22) to the corresponding untreated control as determined by qRT-PCR is shown. GAPDH served as housekeeping gene. Mean ± SEM. n=4 independent donors. **B)** Polarization of CD4^+^ T cells after treatment with the five mediators over 10 days were further analysed by a multiplex ELISA assaying the secretion of IFNγ (Th1/Tc1), TNFα (Th1/Tc1, Th22/Tc22), IL-2 (Th1/Tc1, Th2/Tc2), IL-4 (Th2/Tc2), IL-13 (Th2/Tc2, Th22/Tc22), IL-17a (Th17/Tc17), and IL-22 (Th17/Tc17, Th22/Tc22). Mean ± SEM. n=2 independent donors.

To verify the qPCR results, we conducted a multiplex assay quantifying the cytokines that were released by primary human CD4^+^ and CD8^+^ T cells following exposure to the five mediators (**Fig. 8B, Fig. S7, S8**). Notably, stimulation of CD4^+^ T cells with CD44 did not induce any notable cytokine production. In contrast, C3, FN, SD-4, and TSP-1 stimulated INFγ, IL-2, and TNFα and less pronounced IL-17A production confirming the Th1 polarizing/expanding effects as derived from the qPCR data. While the qPCR data for CD8^+^ T cells did not indicate any effects, the multiplex ELISA showed revealed slightly increased IL4, IL17A, and IL22 production (**Fig. S8**).

**Figure 8.**
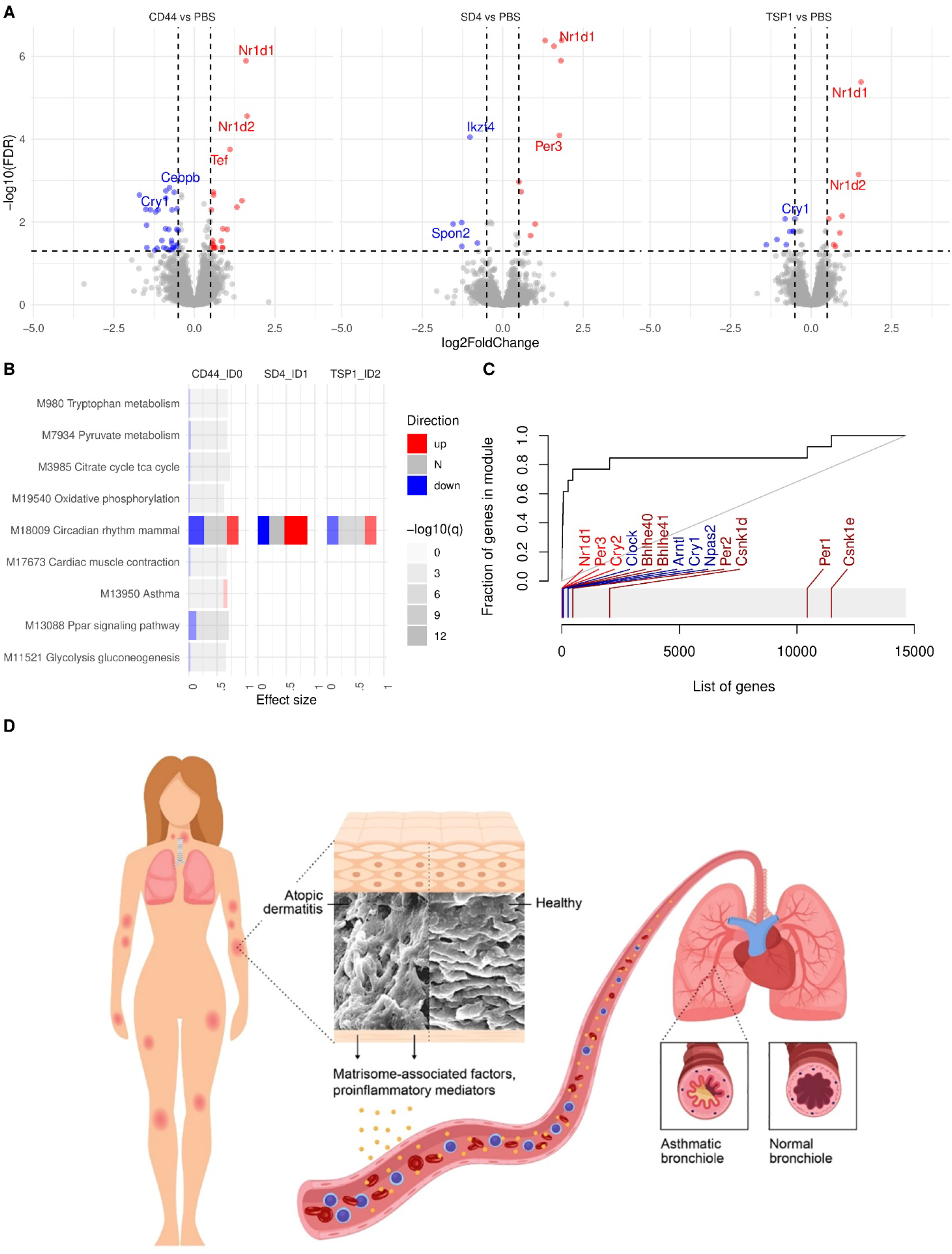
Transcriptomic changes induced by treatment of murine lungs with ECM compounds. **A)** Volcano plots depicting DE genes from exposure of murine lungs to one of three compounds identified in earlier experiments: CD44 vs PBS, SD-4 vs PBS, and TSP-1 vs PBS. Genes coloured red or green were found to be significantly up or down regulated in the treatment group (vs. a PBS control group), respectively. **B)** Results of the gene set enrichment analysis for the KEGG pathways. Columns correspond to contrasts and rows correspond to KEGG pathways. Length of each bar corresponds to the effect size (area under curve, AUC); intensity of the color corresponds to the FDR; red and blue fragments indicate fractions of the pathway which have the absolute log2 fold change larger than 0.5. **C)** Receiver-operator characteristic (ROC) curve showing gene set enrichment for the KEGG pathway “Circadian rhythm mammal” for the contrast SD4 vs PBS. Genes shown in bright colors are significantly up- or down-regulated in the SD4 animals as compared to PBS; red indicates up-regulation, blue indicates down-regulation. **D)** Schematic drawing of the proposed skin-lung crosstalk as potential contributor to the progression of atopic diseases.

Finally, we conducted a transcriptomic profiling of the murine lungs revealing 17-46 significantly DE genes in each treatment group (**Data Set S5**), with the most DE genes in the CD44 treated mice. Strikingly, amongst all three treatment groups, many of the exact same genes were differentially expressed. This includes classic circadian clock genes such as the *nuclear receptor 1d1 (Nr1d1)* and *1d2* (*Nr1d2*) as well as their down-stream regulators *Tef* and *Dbp* (**Fig. 8A**). In contrast, C*ry1, a*nother central circadian clock gene, was significantly down-regulated in CD44-and TSP-1-treated mice.

To functionally characterize the DE genes for the three mediators tested, we applied gene set enrichment analysis to the REACTOME and KEGG pathways (**Fig. 8B, C, Table S4; Fig. S10-S15**). Although significantly enriched pathways were found for all mediators, the majority of the significant enrichments was found for the effect of the CD44 with the REACTOME and KEGG pathways showing congruent enrichments. For CD44, both REACTOME and KEGG indicated enrichment in several gene sets related to asthma, citric acid cycle, and lipid metabolism. Notably, the KEGG pathway “Circadian rhythm” was enriched in all three comparisons (**Fig. 8B, C**; FDR < 10-6 for each of the three comparisons), and that the REACTOME pathway “Circadian clock” was enriched for both CD44 and SD4 mediators (FDR < 0.001 for both comparisons). This enrichment was driven – amongst others – by the genes Nr1d1, Per3, Cry2 (significantly up-regulated in the treated animals) and Clock (significantly down-regulated in the treated animals).

## Discussion

The underlying mechanisms of the atopic march are largely unknown. Here, we proposed the hypothesis of skin-lung crosstalk driven by ECM remodeling, which may promote the progression of atopic diseases. The impact of inter-tissue communication has received increasing attention over the past decade unveiling important interdependencies in health and disease for example between the gut and the lung [*32*] and the gut and brain [*33*] prompting the question as to what other tissue-axes may exist.

For our study, we leveraged an atopic-like skin disease model based on a *FLG* knockdown supplemented with IL-4/IL-13. This model closely emulates hallmarks of AD such as hyperproliferation, elevated skin surface pH, increased TSLP expression, and disturbed skin barrier function [*34, 35*]. As the atopic march originates from skin manifestations, one speculates that skin-derived mediators harness the onset of inflammation in other epithelia. Indeed, here we showed indirect co-cultures of normal bronchial epithelial models with atopic-like skin models triggered an asthma-like phenotype (**Fig. 1C**). The latter was characterized by hyperproliferation, increased mucus production, and up-regulation of classic asthma markers such as TSLP, tenascin, and αSMA [*36*]. Interestingly, the expression of the putatively anti-inflammatory protein, uteroglobin, was increased after the indirect co-culture, potentially as a protective measure [*37*]. Concordant characteristics with respect to αSMA and mucus production were observed in tissue samples from asthmatics and air-liquid interface cultures generated from asthmatic patient-derived cells (**Fig. 1B**).

To identify the causative factors for these effects, a targeted proteomics analysis was conducted identifying fibronectin (FN), TSP-1, C3, CD44, and SD-4 as potentially relevant. The exposure of these mediators to normal bronchial epithelial models triggered similar effects to those found by exposure to the culture medium of atopic-like skin models (**Fig. 4**), this corroborating their contribution to the disease phenotype shown in **Fig. 1**. Previous studies reported correlations between disease activity and severity for these mediators, as well as elevated plasma levels in AD or AA patients [*38-43*]. For example, complement factor C3 can regulate Th2-driven immune responses [*44*], while abolishing C3 expression in mice significantly lowers bronchial hyperreactivity [*45*]. Similarly, FN plasma levels correlate with AD and AA severity [*46*], while shedding of the SD-4 ectodomain into the blood stream can promote AA [*47*]. The hyaluronic acid (HA) receptor CD44 regulates lymphocyte activity through interactions with ECM components such as FN or laminin [*48, 49*]. Also TSP-1 stimulates expression of TGFβ [*50*], a cytokine that facilitates the deposition of FN in asthmatics, thus facilitating airway hyperreactivity and reduced lung function [*51*]. Yet, the actual contribution of these mediators to atopic diseases in general and the atopic march in particular remained largely elusive.

Strikingly, all five mediators have been linked to the skin matrisome, which represents the entirety of skin extracellular matrix (ECM)-associated factors. ECM glycoproteins, collagens, and proteoglycans are considered to constitute the core matrisome, whereas all ECM-affiliated proteins, ECM regulators and secreted factors are considered matrisome-associated [*52*]. FN and TSP-1 belong to the core skin matrisome, SD-4 is a matrisome-associated protein, CD44 is a receptor of ECM proteins such as hyaluronic acid, collagens, and matrix metalloproteinases [*53*], and keratinocyte-or fibroblast-derived C3 is bound by ECM [*54*].

The ECM is a complex structure that provides tissue boundaries and biomechanical properties. Previously, its contribution to tissue homeostasis has been underappreciated, yet, recent data show that the ECM governs tissue homeostasis through biomechanical cues and control over cell survival, proliferation, and differentiation [*52*], as well as inflammatory processes [*55*]. A prominent example is inflammatory bowel disease [*56, 57*] where ECM remodeling has been linked to disease progression and severity [*55, 58-60*]. The concept of ECM remodeling in the context of AD is largely unexplored, which prompted us to investigate the ECM in atopic conditions in more detail.

Proteomic profiling of ECM samples showed a large number of differentially expressed proteins in atopic *versus* non-atopic ECM samples. For example, tenascin, a binding partner of FN [*61*], was significantly more abundant in normal ECM. One can speculate that less tenascin is available to bind FN in atopic samples triggering its release which might lead to pro-inflammatory effects at other locations (**Fig. 4**). However, further investigations are warranted to substantiate this claim. Tenascin also stimulates collagen type I production and reduces the expression of matrix metalloproteinase-1 [*62*]. Hence, its low levels in atopic ECM may also contribute to the structural differences observed by SEM (**Fig. 5A**). In contrast, HTRA1, a serine protease, that facilitates ECM remodeling and fragmentates FN was significantly more expressed in atopic ECM. FN fragments upregulate matrix metalloproteinases which further degrade or remodel ECM [*63, 64*]. The signal transducer and activator of transcription 1-alpha/beta (STAT1) is another interesting protein as it is a pivotal down-stream regulator of the JAK-STAT and TSLP pathway in AD [*65, 66*]. Additionally, extracellular superoxide dismutase (SODE) which acts as a scavenger for reactive oxygen species (ROS) is more abundant in healthy ECM, while low levels facilitate inflammation in atopic skin [*67, 68*].

A complementary transcriptomic analysis of AD patient-derived fibroblasts revealed a significant number of DE genes, which further yielded several AD and atopic march-relevant key pathways including ECM organization and remodeling, cytokine signaling, ECM degradation, and VEGF signaling (**Fig. 6**). Well in line with the previous data, we saw significant changes in matrisome-associated genes such as fibronectin (*FN)* which played a role in almost every analysis that we conducted in the frame of this study, as well as *ITGB5* which codes for a FN receptor. Similarly, *STAT1* and *HSP90AA1* were key nodes within this network, with the former corresponding to the increased STAT1 protein expression in atopic ECM (**Fig. 5**) and the latter coding for the heat shock protein (HSP) 90. HSP90 orchestrates stress responses yet its involvement in inflammatory skin diseases is not fully understood [*69*].

To unravel potential downstream effects on T cells, we exposed primary human T cells to the mediators of interest. All mediators triggered CD4^+^ T cell polarization on gene level as derived from significantly increased mRNA expression of Th1 markers and, although less pronounced, Th17 and Th22 (**Fig. 7A**). These effects were confirmed on protein level showing particularly pronounced effects of TSP-1. In contrast to the *in vitro* data, the immune cell data derived from the *in vivo* experiment were less conclusive, likely due to a rather short administration over only 7-days or the fact that different species were used. It is well established that different T cell subsets contribute to AD in a disease stage-dependent manner [*70*]. Th2 and Th22 cells are predominant in the acute phase, the later also promoting airway inflammation [*20, 71*]. In contrast, Th1 and Th17 cells dominate chronic disease states and correlate with disease activity [*22*]. Although more detailed investigations are warranted, our initial data indicate that skin-derived factors such as TSP-1 and SD-4 exert down-stream effects on T cells. Since the skin harbors tissue-resident T cells and active T cell recruitment into atopic skin occurs, we speculate that these effects may also occur locally *in vivo* triggering un-known down-stream effects that may contribute to the atopic march.

Although we did not observe any functional or histological changes in mice treated with SD-4, TSP-1, or CD44, transcriptomic analyses of the murine lungs showed a dysregulation of central circadian clock genes such as *Nr1d1* and *Nr1d2* (**Fig. 8**). *Nr1d1* codes for REV-ERBa and *Nr1d2* for REV-ERBβ; both are nuclear hormone receptors and core clock genes that are critical regulators of inflammation and metabolism and are involved in the cell-autonomous circadian transcriptional/translational feedback [*72*]. Each cell in our body has components of the cellular circadian clock and undergoes circadian oscillations. This oscillation is driven by an activating arm, *clock* and *bmal1* heterodimer that stimulate the inhibitory arms, Per/Cry and REV-ERBα/REV-ERBβ, which negatively regulate *Bmal1/clock* [*73*]. Notably, in our RNA-Seq analysis (**Fig. 6B**), *TP53* was significantly up-regulated. This is of particular interest as TP53 can repress Per2 expression [*74*]. Another gene that was identified as significantly down-regulated as derived from the RNA-Seq analysis is *HDAC1*. It is well established that class IIa histone deacetylases are regulators of the circadian functions [*75*]. Furthermore, the circadian clock modulates the innate immune response including monocyte and macrophage activation as well as cell trafficking [*76, 77*]. In fact, the role of both REV-ERBα and REV-ERBβ in lung homeostasis and inflammation has just recently been highlighted [*77, 78*] as such that a functional loss promotes inflammation and immune cell infiltration. Concordantly, stabilization or activation through agonists abolish inflammation highlighting their role as putative anti-inflammatory targets. In our study, we found a significant upregulation of *Nr1d1, Nr1d2, Tef*, and *Dbp* potentially as a protective counter measure against the putative harmful mediators. We also found significant down-regulation of *Cry-1* in CD-44 and TSP-1 treated mice indicative of a circadian clock imbalance. One can speculate that this potentially protective measure is abolished at some point causing a disruption of the circadian rhythm in lung tissue and, thus promoting the manifestation of asthma. This finding is of particular interest as a disrupted expression of circadian clock genes in asthmatic individuals is evident [*79*].

Taken together, in this study we detailed the hypothesis of skin-lung crosstalk driven by ECM remodeling as a putative contributor to the progression of atopic diseases (**Fig. 9**). Our data provide initial insights into the presumptive role of skin-derived mediators as contributors to the atopic march and raise important follow-up questions. For example, what triggers the ECM remodeling? Possibly it is the chronic inflammation in AD, or distinct proinflammatory fibroblast subsets that have been recently identified in AD [*80*]. Interestingly, the single cell RNA-Seq study of He et al., described a *COL6A5*^*+*^ *COL18A1*^*+*^ fibroblast subpopulation in AD patients and it was hypothesized that COL18A1 may contribute to ECM disorganization and instability in AD. Conversely, in our bulk RNA-Seq study, *COL18A1* was 5.56-fold significantly downregulated (**Data Set S3**, Fig. 6A), whereas *COL6A5* was not detected, indicating that most cells showed the opposite trends to this rare (<5%) fibroblast population. Future studies should also examine if the release of ECM-derived mediators correlates with disease activity, and if so, could that serve as prognostic or diagnostic marker or for patient stratification? Another very interesting aspect is that *in vivo* the progression from AD to AA takes between 5-8 years. One could speculate that although the plasma levels of the mediators of interest are increased, the levels are likely insufficient to trigger acute effects. What eventually tips the scale towards AA manifestation is unclear. Finally, future studies examining the effect of the mediators in other atopy-relevant epithelia such as the intestine or the nasal epithelium are warranted.

Limitations of our study are that we focused on five mediators; yet the effects induced by the culture medium collected from atopic-like skin models differed from the reactions to the single mediators. This indicates that the culture medium contains other mediators that may be relevant or that the mediator cocktail may be important. Further, although we present AD patient-related data, the samples were accessed retrospectively through a biobank and very limited clinical information was available. Thus, larger prospective studies ideally in high-risk atopic children, are required to assess the plasma profiles of ECM mediators relative to disease severity and activity.

## Materials and Methods

### Cells and tissues

Normal human bronchial epithelial cells (NHBE) and lung fibroblasts (NHLFb) were purchased from Epithelix (Geneva, Switzerland). NHBE were cultured in PneumaCult Ex Plus (STEMCELL Technologies, Vancouver, Canada), NHLFb in fibroblast growth medium (Dulbecco’s Modified Eagle Medium, 10 % fetal calf serum, 5 % L-Glutamine (Sigma-Aldrich, Darmstadt, Germany). Primary human keratinocytes (KC) and fibroblasts (Fb) were isolated from juvenile foreskin according to established standard procedures (with informed consent and ethical approval EA1/081/13). KCs were cultivated in EpiLife supplemented with HKGS (Thermo Scientific, Waltham, MA), Fb were cultivated in fibroblast growth medium (FGM) consisting of DMEM (Sigma-Aldrich, Munich), 10% FBS Superior (Biochrom, Berlin), 2mM L-glutamine (Biochrom, Berlin), 2 mM, Penicillin/Streptomycin 400 U/ml/400 μg/ml (Biochrom, Berlin) containing. Blood plasma samples from atopic patients and non-atopic controls were obtained through the BC Children’s Hospital biobank (CREB approval #H19-00446). Fibroblasts from AD patients were obtained from plucked scalp hair follicles as described elsewhere (age between 20 and 30, with ethical approval and written consent, EA1/345/14) [*81, 82*]. No differences between fibroblasts isolated from skin biopsies and plucked scalp hair follicles as a non-invasive cell source have been previously demonstrated [*82*].

De-identified asthmatic and non-asthmatic donor lungs, not suitable for transplantation and donated to medical research, were obtained though the International Institute for the Advancement of Medicine (Edison, NJ, USA). Epithelial cells were isolated by protease digestion, as previously described [*83*]. Donor deaths were primarily attributable to head trauma in non-asthmatic patients, whereas five of six patients with asthma were thought to have died during exacerbations of their asthma. This study was approved by the Ethics Committee of the University of British Columbia. Donor airways and donor-matched ALI epithelial cultures were generated using cells at Passages 1 or 2, and analyzed using immunohistochemistry

### Tissue models and co-culture experiments

#### Atopic-like skin disease equivalents

Normal (*FLG+*) and diseased (*FLG-*_IL4-/IL-13_) skin equivalents were generated according to previously published procedures [*34, 35*] (see Supplementary Materials for details). Briefly, skin were equivalents generated from primary human dermal fibroblasts (FB) and keratinocytes (KC). FB’s were mixed with bovine collagen type I and FBS and brought to neutral pH and poured into 6-well inserts. After 4 h of collagen solidification, KCs (with or without FLG knock-down) were seeded on top of the dermal matrix. After 24 h skin model was air-lifted and medium was changed to a differentiation medium with medium change every second day.

#### Bronchial epithelial models

For the generation of the bronchial epithelial models, 6.4×10^4^ NHLFb were mixed with FBS, bovine collagen I (PureCol, Advanced BioMatrix, San Diego, USA) and brought to a neutral pH. This mixture was poured into 12-well cell culture inserts (growth area 0.9 cm^2^) and incubated for 2 h at 37°C. Subsequently, 9×10^5^ NHBE were added on top of the collagen matrix. The next day, bronchial epithelial equivalents were lifted to the air-liquid interface and medium was changed to differentiation medium (PneumaCult ALI, Stem Cell, Vancouver, BC, Canada). Medium change was performed every other day for 21 days.

#### Generation of self-assembled ECM with patient-derived (AD) fibroblasts

Fibroblasts from AD patients and skin-healthy volunteers (age between 20-30; with ethical approval, EA1/345/14) were isolated and cultured according to previously published procedures [*82*]. To trigger ECM self-assembly, 2,800 fibroblasts were cultured in 12-well (0.4 um pore size) in low glucose DMEM media supplemented with 10% FBS and 1%Pen/Strep. Media was changed every other day and the ECMs were harvested after 28 days.

### Indirect Skin-Lung Co-Cultures

To study the effects of atopic skin on normal bronchial epithelial models, medium from healthy and diseased (*FLG-*_IL4-/IL-13_) skin equivalents was mixed 1:1 with fresh PneumaCult ALI medium and added to bronchial epithelial models for six days with medium change every second day. To avoid data skewing through cytokine supplementation, the skin disease models were starved from IL4/IL13 at least for 24h prior to its addition to bronchial epithelial models.

To investigate the effects of atopic-like skin-derived mediators on the bronchial epithelial models, 50 ng/ml C3, 1 ug/ml fibronectin (FN), 5 ng/ml syndecan-4 (SD4), 50 ng/ml CD44, or 5 ng/ml thrombos-pondin-1 (TSP-1) (R&D Systems, Abingdon, United Kingdom) were directly added to the culture medium at day 15 of the tissue culture. At day 21, the culture medium was collected and bronchial epithelial models were harvested by fixation in 4% PFA for 24 h and subsequent embedding into paraffin for subsequent processes as (immune-)histological analysis, RT qPCR, and ELISA.

### Histology and immunofluorescence staining

Hematoxylin and eosin (H&E) staining, Alcian blue staining to visualize the mucus on the bronchial epithelial models, and immunofluorescence were performed according to standard protocols (for details, see supplementary material). Antibodies used for immunofluorescence are listed in Supplementary Table S1.

### RT qPCR and western blot

To analyse relevant gene and protein expression levels in bronchial epithelial models, mRNA and proteins were isolated using the AllPrep DNA/RNA/Protein Mini Kit (Quiagen, Hilden, Düsseldorf) according to the manufacturer’s instructions. mRNA isolation from T cells was performed using the innuPREP RNA mini 2.0 kit (Analytik Jena, Berlin, Germany). For cDNA synthesis, the iScript™ cDNA Synthesis Kit (Bio-Rad Laboratories, Munich, Germany) was used. The subsequent RT-qPCR was performed using SYBR Green I Master Mix (Bio-Rad Laboratories, Munich, Germany). The primer sequences are listed in supplementary **Table S2**. Glyceraldehyde-3-phosphate dehydrogenase (GAPDH) served as housekeeping gene.

For western blot analysis, the protein amount was quantified by a BCA Protein Assay (Thermo Scientific, Waltham, MA). Subsequent western blot analysis was performed according to standard protocols (see supplementary material). Blots were cut horizontally in three parts for protein staining (**Fig. S9**). The antibody list can be found in supplementary material (**Table S1**). Protein expression levels were semi-quantified by densitometry and normalized to β-actin or GAPDH levels using ImageJ version 1.46r (National Institutes of Health, Bethesda, MD, USA).

### ELISA

Released cytokine levels were quantified using commercial ELISA kits according to the manufacturer’s instructions. Elisa kits for TSLP, IL-6, IL-8, and IL-33 were purchased from Thermo Fisher (Thermo Fisher Scientific, Waltham, MA, USA) and for desmosine from My BioSource, San Diego, USA. ELISA DuoSet kits for IL-25, fibronectin, syndecan-4, CD44, and thrombospondin-1 were purchased from R&D Systems (Abingdon, United Kingdom). To quantify C3, a colorimetric ELISA kit was purchased from R&D Systems (Abingdon, United Kingdom).

### Collagen quantification

To quantify the collagen content in ECM, a colorimetric hydroxyproline assay kit was used according to manufacturer’s instructions (Abcam, Cambridge, UK). Briefly, the ECM was hydrolyzed with concentrated NaOH at 120°C for 1 h. 10 N concentrated HCl was added to neutralize residual NaOH and the supernatant was collected after centrifugation. Samples hydrolysate were evaporated at 65°C and dissolved in an oxidation reagent mix. Finally, absorbance was measured at 560 nm on a microplate reader.

### Scanning Electron Microscopy (SEM)

SEM was utilized to visualize the fiber arrangement and structure of ECM produced by AD patient-derived- and normal fibroblasts as well as excised human skin (#CREB H19-03096). First, the ECM were carefully cut under a dissection microscope and subsequently fixed with 2.5% glutaraldehyde (pH 7.4) at room temperature for 10 min. Samples were then dehydrated stepwise by immersing them into increasing concentrations of ethanol. Subsequently, the samples were transferred into a critical point dryer after which they were mounted carefully in lateral view on a metal stub with double sided carbon adhesive tape that has been previously coated with Au/Pd using in Leica EM MED020 Coating System. Samples were then imaged by a Helios NanoLabTM 650 (FEI, Hillsboro, OR) scanning electron microscope, operated at 2–10 kV.

### Proteome analysis

#### Secretome analysis of skin disease equivalents

In preparation for the secretome analysis, fetal calf serum (FBS) was removed from culture medium of the skin equivalents at day 10, followed by medium collection at day 12 and day 14. The total protein concentration in the medium was then determined with a Pierce™ BCA Protein Assay Kit (Thermo Scientific, Waltham, Massachusetts, USA), followed by sample mixture (2:1) with standard SDS-polyacrylamide gel electrophoresis (PAGE) sample buffer (supplemented with 100 mM DTT), boiling at 95°C for 5 min, and subsequently the application of 30 μg protein onto SDS-PAGE (8% bis-acrylamid running gel). The gel was stained with SimplyBlue™ SafeStain according to manufacturer’s instructions (Invitrogen, Carlsbad, CA). Following SDS-PAGE, bands of interest were excised and cut into 1-2mm pieces and prepared for MS analysis. For full details, see supplementary materials.

#### Semi-quantitative proteomic analysis of ECM

Each ECM sample was lysed with 50% TFE protocol (30uL TFE:30uL H2O), sonicate for 5 min, freezed, thawed, sonicated for 10 min, and pH adjusted by 1 M Tris pH 8.5. Then taken from 26 μg to 86 μg of each sample for further process. Reduction of disulfide bonds of proteins was done by tris(2-carboxyethyl)phosphine (TCEP) solution (1μg to every 50 μg protein) and incubated for 20 min at room temperature. Blocking free sulfhydryl groups was performed by chloroacetamide (CAA) solution (5 μg to every 50 ug protein) and incubated for 10 min at 95°C in the dark. Then samples were diluted by four volumes of 50 mM ammonium bicarbonate (pH ∼ 8) for pH correction, followed by adding LysC enzyme (1 μg to every 50 μg protein) and incubated 2 h at 37°C. Lastly, samples were digested with Trypsin (1 μg to every 50 μg protein) for 19 h at 37°C, followed by the second round of Trypsin digestion (1 μg to every 125 μg protein), incubated 5 h at 37C. Trypsin activity was quenched by acidifying samples by adding 1% TFA for a final pH < 2.5 of the samples followed by cleaned up via STAGE-Tip purification as described above. MS analysis was then performed as described above.

The acquired data were subsequently searched by MaxQuant (V1.6.10.43) against the Uniprot protein database for Homo sapiens, and LFQ intensities extracted and normalized using the MaxLFQ algorithm [*84*], with 20 ppm and 30 ppm mass accuracies for precursor and product ion masses, respectively, and a 1% false discovery rate cut-off. Label-free quantitation results were then filtered and tested with statistical software as follow. Data were loaded into the Perseus computational Proteomics Platform (V1.6.15.0) [*85*]. First, the data were filtered for common contaminants and false-positive identification, then log-transformed, followed by checking for tightness of technical replicates (Pearson correlation > 0.95).

### RNA-Sequencing and bioinformatic analysis

First, bulk RNA was isolated from AD patient-derived fibroblasts and healthy control samples using the Invitrogen PureLink RNA mini kit (Thermo Scientific, Burnaby, Canada) according to the manufacturer’s protocol. Sample quality control was then performed using the Agilent 2100 Bioanalyzer. Qualifying samples were then prepped following the standard protocol for the NEBnext Ultra ii Stranded mRNA (New England Biolabs). Sequencing was performed on the Illumina NextSeq 500 with Paired End 42bp×42bp reads with a sequencing depth of 20 million paired end reads per sample. Sequencing data was demultiplexed using Illumina’s bcl2fastq2. De-multiplexed read sequences were then aligned to the Homo sapiens/hg19 reference sequence using STAR aligner [*86*]. Assembly and differential expression were estimated using Cufflinks (http://cole-trapnell-lab.github.io/cufflinks/) through bioinformatic apps available on the Illumina Sequence Hub. There were a minimum of 13,978,753, median of 17,760,215, and a maximum of 23,957,122 uniquely mapped reads. Differential expression (DE) analysis was performed using DESeq2 v1.28.1 [*87*], and significance for DE genes was determined using an absolute fold change value of ≥ 1.5 and adjusted p-value ≤ 0.05. Since samples were sequenced in multiple runs, batch was included in the DESeq2 design formula to control for potential batch effects. Pathway enrichment of DE genes was performed using Sigora v3.0.5 [*88*], with significance defined as a Bonferroni-corrected p-value ≤ 0.001. Network analysis was done by uploading genes and their respective fold change values to NetworkAnalyst for construction of protein-protein interaction (PPI) networks [*89*].

#### RNA-Seq analysis of murine lungs

Sequence reads were checked for quality with FastQC v0.11.8 and MultiQC v1.8. Sequences were then aligned to the Ensembl mouse genome GRCm39 using the STAR alignment program (v2.7.3a), and counts generated using HTSeq’s count function (v0.11.2). There were a minimum of 2,749,374, median of 3,846,763 and maximum of 5,437,688 uniquley mapped reads. Differential expression (DE) analysis was performed using DESeq2 v1.32 [*87*]. The linear model was fit with treatment and batch as coefficients. P-values were corrected for mul-tiple testing using the Benjamini-Hochberg method [*90*]. Genes were considered significant based on an FDR of < 0.05 and absolute fold change > 1.5. Gene set enrichment was tested using the CERNO algorithm from the R package tmod, version 0.46.2 [*91*] on genes or-dered by decreasing p-value and applied to gene sets from the R package msigdbr, version 7.4.1. These included gene set collections from the databases REACTOME [*92*] and KEGG [*93*].

### T cell experiments

Buffy-coat preparations were purchased from the German Red Cross (DRK-Blutspendedienst Ost, Berlin, Germany; with informed consent and ethical approval EA2/067/15). Naïve human CD4^+^ and CD8^+^ T cells were purified from PBMCs by negative selection using magnetic-activated cell sorting beads according to the manufacturer’s instructions (MACS; Miltenyi-Biotec, Bergisch-Gladbach, Germany). T cells were cultivated with RPMI containing 10% AB-serum and 1% pen/strep. Subsequently, the T cells were activated using CD3/CD28-antibody (BioLegend, San Diego, USA). Here, 96-well round bottom plates were pre-coated with CD3/CD28 for 2 h at 37°C and 50.000 naive T cells were added along with 50 ng/ml C3, 1 μg/ml FN, 5 ng/ml SD4, 50 ng/ml CD44, or 5 ng/ml TSP-1 were added, respectively.

After 48h, the T cells were transferred to CD3/CD28-free wells and stimulated with each compound for 8 more days with medium change every other day. After 12 days, T cells were re-stimulated with PMA/ionomycin (Merck, Darmstadt, Germany) for 6h. A cultivation period over 12 days ensured clear polarization towards one specific subset; thus, avoiding misleading signals of residual subsets that often occur after shorter timepoints. The culture medium was subsequently collected and T cell mRNA was isolated using the innuPREP RNA Mini Kit 2.0. The cell culture medium was then subjected to multiplex ELISA (Merck, Darmstadt, Germany) analysis to quantify IFNγ, TNFα, IL-2, IL-4, IL-13, IL-17a, and IL-22 according to manufacturer’s instruction, each well was measured on a MAG-PIX system with xPONENT 4.2 software.

### *In vivo* experiments

Female and male BALB/c (BALB/cAnNCrl) mice at the age of 5-6 weeks were purchased from Charles River (Sulzfeld, Germany). All mice were kept in groups of three mice per cage (type III makrolon) with a 12 h light/ dark cycle at 22°C. Water and standard diet (Altromin, Lage/Lippe, Germany) were provided ad libitum. The animal experiments have been ethically approved by the LaGeSo, Berlin, Germany (G0204/20). Groups were randomly assigned per cage and people working on implantation of pumps, observation, scoring, organ explantion, data evaluation, and statistical analysis were fully blinded. Mice were acclimatized to their new housing environments for 14 days prior to the experiments. The back of the mice was depilated (Veet depilation crème sensitive skin, RB Healthcare, Hull, UK) three days prior to the surgical procedure. On day 0 of the experiment, Alzet micro-osmotic pumps 1007D (Charles River, Sulzfeld, Germany) were filled with 100 μl PBS only for control group, 84 ng/ 100 μl syndecan-4 in PBS, 21μg / 100 μl thrombospondin-1 in PBS, or 105 μg/100 μl CD44 in PBS directly before implanting. The resulting release is 12 ng/d SD-4, 3 μg/d TSP-1, and 15 μg/d CD44 with a release rate of 0.5 μl/hr. The implantation was performed according to [*94*]. For full details, see supplementary material. On day 4 a 100 μl blood sample was collected. Measurement of the temperature and wound control were performed every day until the mice were sacrificed at day 7. Simultaneously, we isolated the T cells from the mouse spleen following a modified protocol as described previously [*95*] (see supplementary materials).

### Histological analyses of murine lungs

Histopathology was performed as previously described in detail [*96*]. Prior to preparation of the lungs, the trachea was ligated to prevent alveolar collapse. After careful removal from the thoracical cavity, lungs were immersions-fixed in 4 % buffered formalin (pH 7.0) for 24 h, embedded in paraffin and cut in 2 μm sections. Slides were dewaxed in xylene, rehydrated in graded ethanol’s and stained with hematoxylin and eosin (H&E). Three evenly distributed sections per lung were microscopically evaluated in a blinded, randomized fashion by a board-certified veterinary pathologist.

### Transcriptome analyses of murine lungs

#### RNA extraction

For each of the 24 mice, three formalin-fixed, paraffin-embedded whole lung sections of 10 μm thickness were used for extraction of total RNA using the RecoverAll Total Nucleic Acid Isolation Kit for FFPE, Invitrogen (PN: AM1975) following the manufacturer’s guidelines. The 200 μl volume option was used. Samples were DNaseI digested during extraction and subsequently stored at -80 °C.

#### RNA quality control and library preparation

Total RNA concentrations were measured using a UV-Vis spectrophotometer Nanodrop2000c (Thermo Fisher). Quality of total RNA was determined with an Agilent 5300 Fragment Analyzer DNF-471F33 - SS Total RNA 15nt - FFPE Illumina DV200 method mode. Approximately 300 ng of DNase I digested total RNA in 5 μl were processed using the QuantSeq 3’ mRNA-Seq FWD Library Preparation Kit (Lexogen, Vienna). User guide version 015UG009V0251 was followed according to the manufacture’s guidelines. The quality of the libraries was determined with Agilent 5300 Fragment Analyzer DNF-474-33 - HS NGS Fragment 1-6000bp method mode.

#### Sequencing

Samples were pooled in equimolar ratio and library pools were quantified using a Qubit dsDNA HS assay kit (Thermo Fisher). Sequencing was performed on an Illumina NextSeq 500 system with SR75 High Output Kit at Lexogen.

### Statistical analysis

Statistical analysis was performed with GraphPad Prism 9 (Version 9.2.0) software. Values are expressed as means ± SEM from at least three independent experiments. A matched-pair T-test was used for comparison of 2 dependent, parametric groups. A one-way ANOVA was used for comparison of >2 parametric groups.

## Data Availability

All data produced in the present study are available upon reasonable request to the authors following acceptance for publication by peer review.

## Acknowledgement

We extend our gratitude to Zheng Tan for preparing the self-assembled ECM samples, the staff of the BCCHR biobank for their support in accessing samples from atopic children, the BRC sequencing core for their excellent scientific and technical support of the transcriptomic analysis, the UBC proteomics core, especially Jason Rogalski and Renata Moravcova for their excellent technical and scientific support during the proteomic analysis as well as the Institute of Biometry and Clinical Epidemiology at the Charité Berlin for their support of the statistical analysis. Tissue schemes were kindly provided by Dr. Anna Löwa.

## Author contribution

Conceptualization: PG, SH, JW, WB

Methodology: PG, JW, OK, PP, REWH, TMB, LL, AT, AV, JMW, DB

Investigation: PG, JW, OK, PP, TMB, AV, TLH, JMW, DB

Visualization: PG, TLH, TMB, PP, JMW, SH

Funding acquisition: SH, ADG, REWH, PG

Supervision: SH, WB, REWH, ADG

Writing – original draft: PG, SH, TMB, PP, JMW, WB

Writing – review & editing: PG, SH, REWH, JMW, TLH, DB, WB, ADG

## References

1. Silverberg, J.I., S. Barbarot, A. Gadkari, E.L. Simpson, S. Weidinger, P. Mina-Osorio, A.B. Rossi, L. Brignoli, G. Saba, I. Guillemin, M.C. Fenton, S. Auziere, and L. Eckert, Atopic dermatitis in the pediatric population: A cross-sectional, international epidemiologic study. Ann Allergy Asthma Immunol, 2021. 126(4): p. 417-428.e2.

2. Barbarot, S., S. Auziere, A. Gadkari, G. Girolomoni, L. Puig, E.L. Simpson, D.J. Margolis, M. de Bruin-Weller, and L. Eckert, Epidemiology of atopic dermatitis in adults: Results from an international survey. Allergy, 2018. 73(6): p. 1284–1293.

3. Toledo, M.F., B.M. Saraiva-Romanholo, R.C. Oliveira, P.H. Saldiva, L.F. Silva, L.F. Nascimento, and D. Sole, Changes over time in the prevalence of asthma, rhinitis and atopic eczema in adolescents from Taubate, Sao Paulo, Brazil (2005-2012): Relationship with living near a heavily travelled highway. Allergol Immunopathol (Madr), 2016. 44(5): p. 439–44.

4. Aw, M., J. Penn, G.M. Gauvreau, H. Lima, and R. Sehmi, Atopic March: Collegium Internationale Allergologicum Update 2020. Int Arch Allergy Immunol, 2019: p. 1–10.

5. Wright, B.L. and J.M. Spergel, Eosinophilic Esophagitis. J Allergy Clin Immunol Pract, 2018. 6(5): p. 1799–1801.

6. Akdis, C.A., Does the epithelial barrier hypothesis explain the increase in allergy, autoimmunity and other chronic conditions? Nat Rev Immunol, 2021.

7. Tran, M.M., D.L. Lefebvre, C. Dharma, D. Dai, W.Y.W. Lou, P. Subbarao, A.B. Becker, P.J. Mandhane, S.E. Turvey, and M.R. Sears, Predicting the atopic march: Results from the Canadian Healthy Infant Longitudinal Development Study. J Allergy Clin Immunol, 2018. 141(2): p. 601-607.e8.

8. Ricci, G., A. Patrizi, E. Baldi, G. Menna, M. Tabanelli, and M. Masi, Long-term follow-up of atopic dermatitis: retrospective analysis of related risk factors and association with concomitant allergic diseases. J Am Acad Dermatol, 2006. 55(5): p. 765–71.

9. Irvine, A.D., W.H. McLean, and D.Y. Leung, Filaggrin mutations associated with skin and allergic diseases. N Engl J Med, 2011. 365(14): p. 1315–27.

10. Rodriguez, E., H. Baurecht, E. Herberich, S. Wagenpfeil, S.J. Brown, H.J. Cordell, A.D. Irvine, and S. Weidinger, Meta-analysis of filaggrin polymorphisms in eczema and asthma: robust risk factors in atopic disease. J Allergy Clin Immunol, 2009. 123(6): p. 1361-70.e7.

11. Drislane, C. and A.D. Irvine, The role of filaggrin in atopic dermatitis and allergic disease. Ann Allergy Asthma Immunol, 2020. 124(1): p. 36–43.

12. Han, H., F. Roan, and S.F. Ziegler, The atopic march: current insights into skin barrier dysfunction and epithelial cell-derived cytokines. Immunol Rev, 2017. 278(1): p. 116–130.

13. Han, H., W. Xu, M.B. Headley, H.K. Jessup, K.S. Lee, M. Omori, M.R. Comeau, A. Marshak-Rothstein, and S.F. Ziegler, Thymic stromal lymphopoietin (TSLP)-mediated dermal inflammation aggravates experimental asthma. Mucosal Immunol, 2012. 5(3): p. 342–51.

14. Zhang, Z., P. Hener, N. Frossard, S. Kato, D. Metzger, M. Li, and P. Chambon, Thymic stromal lymphopoietin overproduced by keratinocytes in mouse skin aggravates experimental asthma. Proc Natl Acad Sci U S A, 2009. 106(5): p. 1536–41.

15. Demehri, S., M. Morimoto, M.J. Holtzman, and R. Kopan, Skin-derived TSLP triggers progression from epidermal-barrier defects to asthma. PLoS Biol, 2009. 7(5): p. e1000067.

16. Leyva-Castillo, J.M., P. Hener, H. Jiang, and M. Li, TSLP produced by keratinocytes promotes allergen sensitization through skin and thereby triggers atopic march in mice. J Invest Dermatol, 2013. 133(1): p. 154–63.

17. Soumelis, V., P.A. Reche, H. Kanzler, W. Yuan, G. Edward, B. Homey, M. Gilliet, S. Ho, S. Antonenko, A. Lauerma, K. Smith, D. Gorman, S. Zurawski, J. Abrams, S. Menon, T. McClanahan, R. de Waal-Malefyt Rd, F. Bazan, R.A. Kastelein, and Y.J. Liu, Human epithelial cells trigger dendritic cell mediated allergic inflammation by producing TSLP. Nat Immunol, 2002. 3(7): p. 673–80.

18. Omori, M. and S. Ziegler, Induction of IL-4 expression in CD4(+) T cells by thymic stromal lymphopoietin. J Immunol, 2007. 178(3): p. 1396–404.

19. Adhikary, P.P., Z. Tan, B.D.G. Page, and S. Hedtrich, TSLP as druggable target - a silver-lining for atopic diseases? Pharmacol Ther, 2021. 217: p. 107648.

20. Leyva-Castillo, J.M., J. Yoon, and R.S. Geha, IL-22 promotes allergic airway inflammation in epicutaneously sensitized mice. J Allergy Clin Immunol, 2019. 143(2): p. 619-630.e7.

21. Matsumoto, K., K. Iikura, H. Morita, and H. Saito, Barrier dysfunction in the atopic march-how does atopic dermatitis lead to asthma in children? J Allergy Clin Immunol, 2020. 145(6): p. 1551–1553.

22. Weidinger, S., L.A. Beck, T. Bieber, K. Kabashima, and A.D. Irvine, Atopic dermatitis. Nat Rev Dis Primers, 2018. 4(1).

23. Gerber, P.A., B.A. Buhren, H. Schrumpf, B. Homey, A. Zlotnik, and P. Hevezi, The top skin-associated genes: a comparative analysis of human and mouse skin transcriptomes. Biol Chem, 2014. 395(6): p. 577–91.

24. Lowa, A., M. Jevtic, F. Gorreja, and S. Hedtrich, Alternatives to animal testing in basic and preclinical research of atopic dermatitis. Exp Dermatol, 2018. 27(5): p. 476–483.

25. Lee, H.J., N.Y. Yoon, N.R. Lee, M. Jung, D.H. Kim, and E.H. Choi, Topical acidic cream prevents the development of atopic dermatitis- and asthma-like lesions in murine model. Exp Dermatol, 2014. 23(10): p. 736–41.

26. Choi, J., N. Sutaria, Y.S. Roh, Z. Bordeaux, M.P. Alphonse, S.G. Kwatra, and M.M. Kwatra, Translational Relevance of Mouse Models of Atopic Dermatitis. J Clin Med, 2021. 10(4).

27. Kim, D., T. Kobayashi, and K. Nagao, Research Techniques Made Simple: Mouse Models of Atopic Dermatitis. J Invest Dermatol, 2019. 139(5): p. 984-990.e1.

28. Ewald, D.A., S. Noda, M. Oliva, T. Litman, S. Nakajima, X. Li, H. Xu, C.T. Workman, P. Scheipers, N. Svitacheva, T. Labuda, J.G. Krueger, M. Suarez-Farinas, K. Kabashima, and E. Guttman-Yassky, Major differences between human atopic dermatitis and murine models, as determined by using global transcriptomic profiling. J Allergy Clin Immunol, 2017. 139(2): p. 562–571.

29. Mótyán, J.A., F. Tóth, and J. Tőzsér, Research applications of proteolytic enzymes in molecular biology. Biomolecules, 2013. 3(4): p. 923–42.

30. Elbjeirami, W.M., E.O. Yonter, B.C. Starcher, and J.L. West, Enhancing mechanical properties of tissue-engineered constructs via lysyl oxidase crosslinking activity. Journal of Biomedical Materials Research Part A, 2003. 66A(3): p. 513–521.

31. Sabat, R., K. Wolk, L. Loyal, W.D. Döcke, and K. Ghoreschi, T cell pathology in skin inflammation. Semin Immunopathol, 2019. 41(3): p. 359–377.

32. Depner, M., D.H. Taft, P.V. Kirjavainen, K.M. Kalanetra, A.M. Karvonen, S. Peschel, E. Schmausser-Hechfellner, C. Roduit, R. Frei, R. Lauener, A. Divaret-Chauveau, J.-C. Dalphin, J. Riedler, M. Roponen, M. Kabesch, H. Renz, J. Pekkanen, F.M. Farquharson, P. Louis, D.A. Mills, E. von Mutius, J. Genuneit, A. Hyvärinen, S. Illi, L. Laurent, P.I. Pfefferle, B. Schaub, E. von Mutius, M.J. Ege, and P.s. group, Maturation of the gut microbiome during the first year of life contributes to the protective farm effect on childhood asthma. Nature Medicine, 2020. 26(11): p. 1766–1775.

33. Tan, H.-E., A.C. Sisti, H. Jin, M. Vignovich, M. Villavicencio, K.S. Tsang, Y. Goffer, and C.S. Zuker, The gut–brain axis mediates sugar preference. Nature, 2020. 580(7804): p. 511–516.

34. Hönzke, S., L. Wallmeyer, A. Ostrowski, M. Radbruch, L. Mundhenk, M. Schäfer-Korting, and S. Hedtrich, Influence of Th2 Cytokines on the Cornified Envelope, Tight Junction Proteins, and ß-Defensins in Filaggrin-Deficient Skin Equivalents. J Invest Dermatol, 2016. 136(3): p. 631–9.

35. Vávrová, K., D. Henkes, K. Strüver, M. Sochorova, B. Skolova, M.Y. Witting, W. Friess, S. Schreml, R.J. Meier, M. Schäfer-Korting, J.W. Fluhr, and S. Küchler, Filaggrin deficiency leads to impaired lipid profile and altered acidification pathways in a 3D skin construct. J Invest Dermatol, 2014. 134(3): p. 746–53.

36. Kudo, M., Y. Ishigatsubo, and I. Aoki, Pathology of asthma. Frontiers in Microbiology, 2013. 4(263).

37. Ray, R., M. Choi, Z. Zhang, G.A. Silverman, D. Askew, and A.B. Mukherjee, Uteroglobin suppresses SCCA gene expression associated with allergic asthma. J Biol Chem, 2005. 280(11): p. 9761–4.

38. Nakao, M., M. Sugaya, N. Takahashi, S. Otobe, R. Nakajima, T. Oka, M. Kabasawa, H. Suga, S. Morimura, T. Miyagaki, H. Fujita, Y. Asano, and S. Sato, Increased syndecan-4 expression in sera and skin of patients with atopic dermatitis. Arch Dermatol Res, 2016. 308(9): p. 655–660.

39. Abdel Fattah, M., M. El Baz, A. Sherif, and A. Adel, Complement components (C3, C4) as inflammatory markers in asthma. Indian J Pediatr, 2010. 77(7): p. 771–3.

40. Miura, H., Clinical and experimental studies on fibronectin in bronchial asthma. Allergology International, 2003. 52(2): p. 85–95.

41. Vedel-Krogh, S., K.L. Rasmussen, B.G. Nordestgaard, and S.F. Nielsen, Complement C3 and allergic asthma: a cohort study of the general population. Eur Respir J, 2021. 57(2).

42. Huang, S.W. and K.J. Kao, The relationship between plasma thrombospondin level and the clinical course of atopic dermatitis. Allergy Proc, 1993. 14(5): p. 357–61.

43. Man, M., P.M. Elias, W. Man, Y. Wu, L.Y. Bourguignon, K.R. Feingold, and M.Q. Man, The role of CD44 in cutaneous inflammation. Exp Dermatol, 2009. 18(11): p. 962–8.

44. Zhang, X. and J. Köhl, A complex role for complement in allergic asthma. Expert Rev Clin Immunol, 2010. 6(2): p. 269–77.

45. Drouin, S.M., D.B. Corry, J. Kildsgaard, and R.A. Wetsel, Cutting edge: the absence of C3 demonstrates a role for complement in Th2 effector functions in a murine model of pulmonary allergy. J Immunol, 2001. 167(8): p. 4141–5.

46. Ohke, M., S. Tada, M. Kataoka, K. Matsuo, M. Nabe, and M. Harada, Plasma fibronectin in asthmatic patients and its relation to asthma attack. Acta Med Okayama, 2001. 55(2): p. 91–6.

47. Herum, K.M., I.G. Lunde, B. Skrbic, G. Florholmen, D. Behmen, I. Sjaastad, C.R. Carlson, M.F. Gomez, and G. Christensen, Syndecan-4 signaling via NFAT regulates extracellular matrix production and cardiac myofibroblast differentiation in response to mechanical stress. J Mol Cell Cardiol, 2013. 54: p. 73–81.

48. Borland, G., J.A. Ross, and K. Guy, Forms and functions of CD44. Immunology, 1998. 93(2): p. 139–48.

49. Funaro, A., G.C. Spagnoli, M. Momo, W. Knapp, and F. Malavasi, Stimulation of T cells via CD44 requires leukocyte-function-associated antigen interactions and interleukin-2 production. Hum Immunol, 1994. 40(4): p. 267–78.

50. Crawford, S.E., V. Stellmach, J.E. Murphy-Ullrich, S.M. Ribeiro, J. Lawler, R.O. Hynes, G.P. Boivin, and N. Bouck, Thrombospondin-1 is a major activator of TGF-beta1 in vivo. Cell, 1998. 93(7): p. 1159–70.

51. Moir, L.M., J.K. Burgess, and J.L. Black, Transforming growth factor beta 1 increases fibronectin deposition through integrin receptor alpha 5 beta 1 on human airway smooth muscle. J Allergy Clin Immunol, 2008. 121(4): p. 1034-9.e4.

52. Naba, A., K.R. Clauser, H. Ding, C.A. Whittaker, S.A. Carr, and R.O. Hynes, The extracellular matrix: Tools and insights for the “omics” era. Matrix Biol, 2016. 49: p. 10–24.

53. Ponta, H., L. Sherman, and P.A. Herrlich, CD44: from adhesion molecules to signalling regulators. Nat Rev Mol Cell Biol, 2003. 4(1): p. 33–45.

54. Fernandez-Godino, R., K.M. Bujakowska, and E.A. Pierce, Changes in extracellular matrix cause RPE cells to make basal deposits and activate the alternative complement pathway. Human Molecular Genetics, 2018. 27(1): p. 147–159.

55. Shimshoni, E., I. Adir, R. Afik, I. Solomonov, A. Shenoy, M. Adler, L. Puricelli, F. Sabino, S. Savickas, O. Mouhadeb, N. Gluck, S. Fishman, L. Werner, T.M. Salame, D.S. Shouval, C. Varol, U. Auf dem Keller, A. Podestà, T. Geiger, P. Milani, U. Alon, and I. Sagi, Distinct extracellular-matrix remodeling events precede symptoms of inflammation. Matrix Biol, 2021. 96: p. 47–68.

56. Karamanos, N.K., A.D. Theocharis, T. Neill, and R.V. Iozzo, Matrix modeling and remodeling: A biological interplay regulating tissue homeostasis and diseases. Matrix Biol, 2019. 75-76: p. 1–11.

57. Khokha, R., A. Murthy, and A. Weiss, Metalloproteinases and their natural inhibitors in inflammation and immunity. Nat Rev Immunol, 2013. 13(9): p. 649–65.

58. Capuano, A., E. Pivetta, G. Sartori, G. Bosisio, A. Favero, E. Cover, E. Andreuzzi, A. Colombatti, R. Cannizzaro, E. Scanziani, L. Minoli, F. Bucciotti, A.I. Amor Lopez, K. Gaspardo, R. Doliana, M. Mongiat, and P. Spessotto, Abrogation of EMILIN1-β1 integrin interaction promotes experimental colitis and colon carcinogenesis. Matrix Biol, 2019. 83: p. 97–115.

59. Lohr, K., H. Sardana, S. Lee, F. Wu, D.L. Huso, A.R. Hamad, and S. Chakravarti, Extracellular matrix protein lumican regulates inflammation in a mouse model of colitis. Inflamm Bowel Dis, 2012. 18(1): p. 143–51.

60. Castaneda, F.E., B. Walia, M. Vijay-Kumar, N.R. Patel, S. Roser, V.L. Kolachala, M. Rojas, L. Wang, G. Oprea, P. Garg, A.T. Gewirtz, J. Roman, D. Merlin, and S.V. Sitaraman, Targeted deletion of metalloproteinase 9 attenuates experimental colitis in mice: central role of epithelial-derived MMP. Gastroenterology, 2005. 129(6): p. 1991–2008.

61. Chiquet-Ehrismann, R. and M. Chiquet, Tenascins: regulation and putative functions during pathological stress. The Journal of Pathology, 2003. 200(4): p. 488–499.

62. Choi, Y.E., M.J. Song, M. Hara, K. Imanaka-Yoshida, D.H. Lee, J.H. Chung, and S.T. Lee, Effects of Tenascin C on the Integrity of Extracellular Matrix and Skin Aging. Int J Mol Sci, 2020. 21(22).

63. Grau, S., P.J. Richards, B. Kerr, C. Hughes, B. Caterson, A.S. Williams, U. Junker, S.A. Jones, T. Clausen, and M. Ehrmann, The role of human HtrA1 in arthritic disease. J Biol Chem, 2006. 281(10): p. 6124–9.

64. Sabino, F., F.E. Egli, S. Savickas, J. Holstein, D. Kaspar, M. Rollmann, J.N. Kizhakkedathu, T. Pohlemann, H. Smola, and U. Auf dem Keller, Comparative Degradomics of Porcine and Human Wound Exudates Unravels Biomarker Candidates for Assessment of Wound Healing Progression in Trauma Patients. J Invest Dermatol, 2018. 138(2): p. 413–422.

65. Welsch, K., J. Holstein, A. Laurence, and K. Ghoreschi, Targeting JAK/STAT signalling in inflammatory skin diseases with small molecule inhibitors. Eur J Immunol, 2017. 47(7): p. 1096–1107.

66. Bao, L., H. Zhang, and L.S. Chan, The involvement of the JAK-STAT signaling pathway in chronic inflammatory skin disease atopic dermatitis. Jakstat, 2013. 2(3): p. e24137.

67. Agrahari, G., S.K. Sah, C.T. Nguyen, S.S. Choi, H.Y. Kim, and T.Y. Kim, Superoxide Dismutase 3 Inhibits LL-37/KLK-5-Mediated Skin Inflammation through Modulation of EGFR and Associated Inflammatory Cascades. J Invest Dermatol, 2020. 140(3): p. 656-665.e8.

68. Sah, S.K., G. Agrahari, C.T. Nguyen, Y.S. Kim, K.S. Kang, and T.Y. Kim, Enhanced therapeutic effects of human mesenchymal stem cells transduced with superoxide dismutase 3 in a murine atopic dermatitis-like skin inflammation model. Allergy, 2018. 73(12): p. 2364–2376.

69. Scieglinska, D., Z. Krawczyk, D.R. Sojka, and A. Gogler-Pigłowska, Heat shock proteins in the physiology and pathophysiology of epidermal keratinocytes. Cell Stress Chaperones, 2019. 24(6): p. 1027–1044.

70. Gittler, J.K., A. Shemer, M. Suárez-Fariñas, J. Fuentes-Duculan, K.J. Gulewicz, C.Q. Wang, H. Mitsui, I. Cardinale, C. de Guzman Strong, J.G. Krueger, and E. Guttman-Yassky, Progressive activation of T(H)2/T(H)22 cytokines and selective epidermal proteins characterizes acute and chronic atopic dermatitis. J Allergy Clin Immunol, 2012. 130(6): p. 1344–54.

71. Nakagawa, R., H. Yoshida, M. Asakawa, T. Tamiya, N. Inoue, R. Morita, H. Inoue, A. Nakao, and A. Yoshimura, Pyridone 6, a pan-JAK inhibitor, ameliorates allergic skin inflammation of NC/Nga mice via suppression of Th2 and enhancement of Th17. J Immunol, 2011. 187(9): p. 4611–20.

72. Ikeda, R., Y. Tsuchiya, N. Koike, Y. Umemura, H. Inokawa, R. Ono, M. Inoue, Y. Sasawaki, T. Grieten, N. Okubo, K. Ikoma, H. Fujiwara, T. Kubo, and K. Yagita, REV-ERBα and REV-ERBβ function as key factors regulating Mammalian Circadian Output. Scientific Reports, 2019. 9(1): p. 10171.

73. Mohawk, J.A., C.B. Green, and J.S. Takahashi, Central and Peripheral Circadian Clocks in Mammals. Annual Review of Neuroscience, 2012. 35(1): p. 445–462.

74. Miki, T., T. Matsumoto, Z. Zhao, and C.C. Lee, p53 regulates Period2 expression and the circadian clock. Nat Commun, 2013. 4: p. 2444.

75. Fogg, P.C., J.S. O’Neill, T. Dobrzycki, S. Calvert, E.C. Lord, R.L. McIntosh, C.J. Elliott, S.T. Sweeney, M.H. Hastings, and S. Chawla, Class IIa histone deacetylases are conserved regulators of circadian function. J Biol Chem, 2014. 289(49): p. 34341–8.

76. Bellet, M.M., E. Deriu, J.Z. Liu, B. Grimaldi, C. Blaschitz, M. Zeller, R.A. Edwards, S. Sahar, S. Dandekar, P. Baldi, M.D. George, M. Raffatellu, and P. Sassone-Corsi, Circadian clock regulates the host response to &lt;em&gt;Salmonella&lt;/em&gt. Proceedings of the National Academy of Sciences, 2013. 110(24): p. 9897.

77. Gibbs, J.E., J. Blaikley, S. Beesley, L. Matthews, K.D. Simpson, S.H. Boyce, S.N. Farrow, K.J. Else, D. Singh, D.W. Ray, and A.S. Loudon, The nuclear receptor REV-ERBα mediates circadian regulation of innate immunity through selective regulation of inflammatory cytokines. Proc Natl Acad Sci U S A, 2012. 109(2): p. 582–7.

78. Pariollaud, M., J.E. Gibbs, T.W. Hopwood, S. Brown, N. Begley, R. Vonslow, T. Poolman, B. Guo, B. Saer, D.H. Jones, J.P. Tellam, S. Bresciani, N.C. Tomkinson, J. Wojno-Picon, A.W. Cooper, D.A. Daniels, R.P. Trump, D. Grant, W. Zuercher, T.M. Willson, A.S. MacDonald, B. Bolognese, P.L. Podolin, Y. Sanchez, A.S. Loudon, and D.W. Ray, Circadian clock component REV-ERBα controls homeostatic regulation of pulmonary inflammation. J Clin Invest, 2018. 128(6): p. 2281–2296.

79. Chen, H.C., Y.C. Chen, T.N. Wang, W.F. Fang, Y.C. Chang, Y.M. Chen, I.Y. Chen, M.C. Lin, and M.Y. Yang, Disrupted Expression of Circadian Clock Genes in Patients with Bronchial Asthma. J Asthma Allergy, 2021. 14: p. 371–380.

80. He, H., H. Suryawanshi, P. Morozov, J. Gay-Mimbrera, E. Del Duca, H.J. Kim, N. Kameyama, Y. Estrada, E. Der, J.G. Krueger, J. Ruano, T. Tuschl, and E. Guttman-Yassky, Single-cell transcriptome analysis of human skin identifies novel fibroblast subpopulation and enrichment of immune subsets in atopic dermatitis. J Allergy Clin Immunol, 2020. 145(6): p. 1615–1628.

81. Löwa, A., P. Graff, S. Kaessmeyer, and S. Hedtrich, Fibroblasts from atopic dermatitis patients trigger inflammatory processes and hyperproliferation in human skin equivalents. J Eur Acad Dermatol Venereol, 2020. 34(6): p. e262–e265.

82. Löwa, A., A. Vogt, S. Kaessmeyer, and S. Hedtrich, Generation of full-thickness skin equivalents using hair follicle-derived primary human keratinocytes and fibroblasts. J Tissue Eng Regen Med, 2018. 12(4): p. e2134–e2146.

83. Hackett, T.L., F. Shaheen, A. Johnson, S. Wadsworth, D.V. Pechkovsky, D.B. Jacoby, A. Kicic, S.M. Stick, and D.A. Knight, Characterization of side population cells from human airway epithelium. Stem Cells, 2008. 26(10): p. 2576–85.

84. Cox, J., M.Y. Hein, C.A. Luber, I. Paron, N. Nagaraj, and M. Mann, Accurate proteome-wide label-free quantification by delayed normalization and maximal peptide ratio extraction, termed MaxLFQ. Mol Cell Proteomics, 2014. 13(9): p. 2513–26.

85. Tyanova, S., T. Temu, P. Sinitcyn, A. Carlson, M.Y. Hein, T. Geiger, M. Mann, and J. Cox, The Perseus computational platform for comprehensive analysis of (prote)omics data. Nat Methods, 2016. 13(9): p. 731–40.

86. Dobin, A., C.A. Davis, F. Schlesinger, J. Drenkow, C. Zaleski, S. Jha, P. Batut, M. Chaisson, and T.R. Gingeras, STAR: ultrafast universal RNA-seq aligner. Bioinformatics, 2013. 29(1): p. 15–21.

87. Love, M.I., W. Huber, and S. Anders, Moderated estimation of fold change and dispersion for RNA-seq data with DESeq2. Genome Biol, 2014. 15(12): p. 550.

88. Foroushani, A.B., F.S. Brinkman, and D.J. Lynn, Pathway-GPS and SIGORA: identifying relevant pathways based on the over-representation of their gene-pair signatures. PeerJ, 2013. 1: p. e229.

89. Xia, J., E.E. Gill, and R.E. Hancock, NetworkAnalyst for statistical, visual and network-based meta-analysis of gene expression data. Nat Protoc, 2015. 10(6): p. 823–44.

90. Benjamini, Y. and Y. Hochberg, Controlling the False Discovery Rate: A Practical and Powerful Approach to Multiple Testing. Journal of the Royal Statistical Society. Series B (Methodological), 1995. 57(1): p. 289–300.

91. Zyla, J., M. Marczyk, T. Domaszewska, S.H.E. Kaufmann, J. Polanska, and J. Weiner, Gene set enrichment for reproducible science: comparison of CERNO and eight other algorithms. Bioinformatics, 2019. 35(24): p. 5146–5154.

92. Gillespie, M., B. Jassal, R. Stephan, M. Milacic, K. Rothfels, A. Senff-Ribeiro, J. Griss, C. Sevilla, L. Matthews, C. Gong, C. Deng, T. Varusai, E. Ragueneau, Y. Haider, B. May, V. Shamovsky, J. Weiser, T. Brunson, N. Sanati, L. Beckman, X. Shao, A. Fabregat, K. Sidiropoulos, J. Murillo, G. Viteri, J. Cook, S. Shorser, G. Bader, E. Demir, C. Sander, R. Haw, G. Wu, L. Stein, H. Hermjakob, and P. D’Eustachio, The reactome pathway knowledgebase 2022. Nucleic Acids Res, 2022. 50(D1): p. D687–d692.

93. Aoki-Kinoshita, K.F. and M. Kanehisa, Gene annotation and pathway mapping in KEGG. Methods Mol Biol, 2007. 396: p. 71–91.

94. Lu, H., D.A. Howatt, A. Balakrishnan, J.J. Moorleghen, D.L. Rateri, L.A. Cassis, and A. Daugherty, Subcutaneous Angiotensin II Infusion using Osmotic Pumps Induces Aortic Aneurysms in Mice. J Vis Exp, 2015(103).

95. Bäumer, W., B. Sülzle, H. Weigt, V.C. De Vries, M. Hecht, T. Tschernig, and M. Kietzmann, Cilomilast, tacrolimus and rapamycin modulate dendritic cell function in the elicitation phase of allergic contact dermatitis. Br J Dermatol, 2005. 153(1): p. 136–44.

96. Dietert, K., B. Gutbier, S.M. Wienhold, K. Reppe, X. Jiang, L. Yao, C. Chaput, J. Naujoks, M. Brack, A. Kupke, C. Peteranderl, S. Becker, C. von Lachner, N. Baal, H. Slevogt, A.C. Hocke, M. Witzenrath, B. Opitz, S. Herold, H. Hackstein, L.E. Sander, N. Suttorp, and A.D. Gruber, Spectrum of pathogen- and model-specific histopathologies in mouse models of acute pneumonia. PLoS One, 2017. 12(11): p. e0188251.

