## Supplemental Methods and Data for "EXTRACELLULAR MATRIX REMODELING IN ATOPIC DERMATITIS HARNESSES THE ONSET OF AN ASTHMATIC PHENOTYPE AND IS A POTENTIAL CONTRIBUTOR TO THE ATOPIC MARCH"

**Generation of normal and atopic-like skin models**

For generation of skin models, primary human keratinocytes (KC) and primary human fibroblast (FB) isolated from juvenile (<10 years old) foreskin were used (approved by ethics committee; informed consent was obtained). In a first step, a mix of bovine collagen type I (PureCol, Advanced BioMatrix, San Diego, USA) and 10x Hanks balanced salt solution (Thermo Scientific, Berlin, Germany) was brought to neutral pH. Subsequently, a total of 300.000 FBs per model were added and poured into 6-well cell culture inserts with a growth are of 4.2 cm^2^. After 2 h of incubation (37°C, CO_2_ free) medium was added into the well and on top of the collagen a next incubation step (2 h, 37°C, 95% humidity, 5% CO_2_). Subsequently, 4.2x10^6^ KCs were added on top of the collagen matrix. For generation of diseased equivalents KCs were transfected (HiPerFect®; Qiagen, Hilden, Germany) with FLG specific siRNA (Sequence: CAGCUCCAGACAAUCAGGCACUCAU; NM_002016, Invitrogen, Darmstadt, Germany) 24h prior to generation of skin disease equivalents. 24 h after adding human KCs (with or without FLG knockdown), medium was changed to a differentiation medium (KDM) and skin equivalents were lifted to the air-liquid interface with medium change every second day. *FLG* knockdown was stable for at least 14 days (≥ 90% knockdown efficacy) as determined by qPCR and published previously [1, 2].

### **Immunofluorescence staining**

After fixing skin cryo-sections (4% PFA, < 5 min) or de-waxing lung micro-sections slides were washed with PBS (5 min) and permeabilized with 0.5% (v/v) Triton-X in PBS. Subsequent to washing (5 min PBS, 5 min PBS+0.0025% BSA+0.025% Tween 20), a blocking step with goat serum (1/20 in PBS) was performed for 30 min at RT. Afterwards, primary antibody was diluted (Table **S1)** in PBS/BSA/Tween and drops of 400 µl were placed onto Parafilm inlaid into steel boxes. Slides were then placed upside-down onto 400 µl of antibody dilution and kept at 4°C for over night incubation. For secondary antibody only control PBS/BSA/Tween was used for incubation.

The next day slides were washed in PBS/BSA/Tween (3x5 min) and incubated upside-down with corresponding secondary Alexa-Fluor® 488 or Alexa Fluor® 594 antibody (dilution 1/400 anti-mouse or anti-rabbit) for 1 h (RT, dark) on Parafilm. After several washing steps (3x5 min PBS/BSA/Tween, 2x5 min PBS, 1x30 s ddH_2_O) slides were air dried, mounted with one drop of Anti-fading Mounting Medium DAPI, covered with a cover slip and stored at 4°C for up to 2 days before visualizing protein expression with fluorescence microscope (Keyence, Neu-Isenburg, GER).

### **Western blot**

Proteins were isolated by gently peeling off epidermis from skin models, placing into RIPA buffer containing 1% (v/v) Protease Inhibitor and grinded (30 s, 25 Hz) using a TissueLyzer (Qiagen, Hilden, Germany). After 30 min incubation on ice and centrifugation (4°C, 30 min, 13.200 rpm) samples were stored at -80° until further use. For protein isolation from bronchial epithelial models whole model was used and protein was isolated together with mRNA using Allprep mini DNA/RNA/Protein isolation Kit (Qiagen, Hilden, Germany) according to manufacture’s instructions.

Protein amount was quantified using Pierce^®^ BCA Protein Assay Kit (Thermo Scientific, Schwerte, Germany) according to manufacture’s instructions. Subsequently, the desired amount was mixed 2:1 with laemmli buffer (10 parts loading buffer (3x) and 1-part DTT (30x)), boiled (5 min, 95°C), and centrifuged (11.000 rpm, 5 min, 4°C). A total protein amount of 30 µg/ml was separated by SDS polyacrylamide gel electrophoresis (Bio-Rad, Munich, Germany) — with gel concentration depending on protein size of interest — and separated according to their molecular weight. Afterwards gels were blotted (30 min 100 V, 90 min 150 V) onto previously activated nitrocellulose membrane (Bio-Rad, Munich, Germany), washed with TBST (3x5 min), and blocked with 5% (w/v) skimmed milk powder in TBST (1 h, RT). Gels were incubated with primary antibody (table S1) over night (4°C, dark), washed with TBST the next day (3x5 min), incubated with anti-mouse or anti-rabbit horseradish-peroxidase-conjugated secondary antibody (Cell Signaling, Frankfurt/Main, Germany) (1 h, RT, dark), and washed with TBST (3x5min). Blots were visualized with a PXi/PXi Touch gel imaging system (Syngene, Cambridge, UK) after developing with SignalFire™ ECL reagent or SignalFire™ ECL Elite reagent (Cell Signaling, Frankfurt/Main, Germany). Intensity of bands was relatively quantified with „ImageJ“ (Version 1.46, Wayne Rusband, National Institute for Health, USA) according to respective loading control.

**Proteom analysis**

*Secretome analysis of skin disease equivalents:* The bands of interest were then washed with de-stain buffer (50cmM NH_4_HCO_3_/100% EtOH – 6:4) and subsequently dehydrated in ethanol. Reduction of disulfide bonds was achieved by incubation with 10 mM DTT for 45 min at 56°C, followed by alkylation in 55 mM IAA for 30 min at RT in the dark. Gel pieces were then dehydrated in EtOH, rehydrated in digestion buffer (50 nM NH_4_HCO_3_ – pH 8), dehydrated again, and then the digestion was done in digestion buffer, incubated overnight at 37°C. Digestion was stopped with 1% TFA, and samples extracted twice with extraction solution (acidified water with acetonitrile – 40% ACN, 0.1% TFA). Samples were then concentrated via vacuum centrifugation. Subsequently, extracted peptide samples were then cleaned up via STAGE-tip purification. In brief, re-solubilized acidified sample was forced through conditioned and equilibrated column with 7 mm of C18 packing, washed with 1% TFA twice, and eluted into clean tubes by buffer containing 40% ACN, 0.1% TFA, then dried down.

For MS analysis, the samples were reconstituted in 2% ACN, 0.5% formic acid, and the peptides were analyzed using a quadrupole – time of flight mass spectrometer (Impact II; Bruker Daltonics) on-line coupled to an EasyLC 1000 HPLC (ThermoFisher Scientific) using a Captive spray nanospray ionization source (Bruker Daltonics) including Aurora Series Gen2 (CSI) analytical column, (25 cm x 75 μm 1.6 μm FSC C18, with Gen2 nanoZero and CSI fitting; Ion Opticks, Parkville, Victoria, Australia), and a μ-pre-column, 300 μm ID x 5 mm, C18 PepMap, 5 μm, 100 A, Thermo Scientific, Waltham, Massachusetts, US).

***In vivo* mouse experiment**

Female and male BALB/c (BALB/cAnNCrl) mice at the age of 5-6 weeks were purchased from Charles River (Sulzfeld, Germany). All mice were kept in groups of three mice per cage (type III makrolon) with a 12 h light/ dark cycle at 22°C. Water and standard diet (Altromin, Lage/Lippe, Germany) were provided ad libitum. The animal experiments have been ethically approved by the LaGeSo, Berlin, Germany (G0204/20). Groups were randomly assigned per cage and people working on implantation of pumps, observation, scoring, organ explantion, data evaluation, and statistical analysis were fully blinded. Pumps were loaded by one person and labeled with numbers representing the identity of each mouse.

Mice were acclimatized to their new housing environments for 14 days prior to the experiments. The back of the mice was depilated (Veet depilation crème sensitive skin, RB Healthcare, Hull, UK) three days prior to the surgical procedure. On day 0 of the experiment, Alzet micro-osmotic pumps 1007D (Charles River, Sulzfeld, Germany) were filled with 100 µl PBS only for control group, 84 ng/ 100 µl syndecan-4 in PBS, 21µg / 100 µl thrombospondin-1 in PBS, or 105 µg/100 µl CD44 in PBS directly before implanting. The resulting release is 12 ng/d SD-4, 3 µg/d TSP-1, and 15 µg/d CD44 with a release rate of 0.5 µl/hr. The implantation was performed according to [3].

Anesthesia was induced with 3 % isoflurane (Isofluran CP®, CP Pharma Handelsgesellschaft mbH, Burgdorf, Germany) in 100% oxygen in an anesthetic chamber with sliding cover (Evonik Plexiglas, 240 x 140 x 120 mm). The chamber was not prefilled to prevent distress. After surgical tolerance began, mice were transferred to a nose cone and anesthesia was maintained with 1.5 % isoflurane in 100 % oxygen. Mice were placed in an abdominal position on a heating pad (36 °C). Eye ointment (Vitagel®, Bausch + Lomb GmbH, Berlin, Germany**)** was administered to both eyes to prevent the eyes from drying out. 2 % lidocaine hydrochloride (bela-pharm GmbH & Co. KG, Vechta Germany) was injected subcutaneously (s. c.) with an insulin syringe (BD Micro-Fine™ +, BD Medical – Diabetes Care, Le Pont de Claix Cedex, France) at the incision site. Meloxicam (Metacam 2mg/ml, Böhringer Ingelheim Vetmedica GmbH, Ingelheim/Rhein, Germany) was administered 1 h prior and 12 and 24 h after the surgical procedure at a concentration of 5 mg/kg (s.c.).

The shaved area was swapped with Braunol® (B. Braun Deutschland GmbH & Co. KG, Melsungen, Germany) followed by three wipes of 70 % ethanol (Carl Roth GmbH + Co. KG, Karlsruhe, Deutschland). Behind the shoulder blades a 0.5 - 1 cm incision, perpendicular to the tail, was made with a surgical scalpel. With a hemostat a pouch for the Alzet pump was created under the skin. A bolus of approximately 0,5 - 1 ml of sterile 0.9 % NaCl (B. Braun Deutschland GmbH & Co. KG, Melsungen, Germany) was applied into the pocket. Afterwards the Alzet pump was inserted into the pouch. The incision sites were pinched together with two hemostats and the wound was closed with 1 – 2 wound clips (11 x 2 mm; AESCULAP, B. Braun Deutschland GmbH & Co. KG, Melsungen, Germany). The whole implantation process took about 5 minutes per mouse. Afterwards, mice were placed in a dark cage on a heating pad for the recovery period. On day 4 a 100 µl blood sample was collected from V. facialis with a 5 mm Goldenrod™ Animal Lancet (Medipoint Inc. Mineola, NY, USA). Measurement of the temperature and wound control were performed every day until the mice were sacrificed at day 7. For sample collection mice were deeply anesthetized with an intraperitoneal injection of ketamine-xylazine-NaCl mix (100 µl/ 10 g bodyweight). The mix contained two parts of Ketamin 10 % 100 mg/ml (bela-pharm GmbH & Co. KG, Vechta Germany), 1-part xylazine 20 mg/ml (Xylariem®, Ecuphar N.V., Oostkamp, Belgium) and nine parts sterile NaCl. After loss of all reflexes abdominal skin and cavity were opened and mice were exsanguinated via the V. cava with a 24G needle. Spleen, thymus, lung and intestines were removed for further investigations.

Simultaneously, we isolated the T cells from the mouse spleen following a modified protocol as described previously [4]. Briefly, the spleen was squeezed through a cell strainer, which resulted in a single-cell suspension in the filtrate. These cells were incubated in an erythrocyte lysis buffer for 5 min followed several washing steps. Subsequently, CD4^+^ T cells were isolated by negative selection using magnetic-activated cell sorting beads according to the manufacturer’s instructions (MACS; Miltenyi-Biotec, Bergisch-Gladbach, Germany). Finally, mRNA from CD4^+^ T cells was isolated as mentioned above.

**Tables**

**Table S1.** Antibodies used in immunofluorescence (IF) and western blot (WB)

| Antibody | IF | WB | specimen | clone | isotype |
| --- | --- | --- | --- | --- | --- |
| **⍺-acet. Tubulin** | 1:2,000 |  | mouse | monoclonal (6-11B-1) | IgG2b |
| **⍺-Smooth muscle actin** | 1:200 | 1:1,000 | rabbit | polyclonal | IgG |
| β-Actin |  | 1:1,000 | mouse | monoclonal (15G5A11/E2) | IgG1 |
| E-Cadherin | 1:100 |  | mouse | monoclonal (67A4.2.1) | IgG1 |
| Filaggrin | 1:1,000 | 1:1,000 | mouse | polyclonal | IgG |
| GAPDH |  | 1:1,000 | rabbit | monoclonal (14C10) | IgG |
| Ki-67 | 1:200 |  | mouse | monoclonal (Ki-67P) | IgG |
| MUC5AC | 1:600 | 1:100 | mouse | monoclonal (CLH2) | IgG1 |
| PAR-2 | 1:200 | 1:200 | rabbit | monoclonal (SAM11) | IgG2A |
| SCCA-1 | 1:125 | 1:1,000 | rabbit | monoclonal (886524) | IgG2B |
| sPLA-2 | 1:400 | 1:1,000 | rabbit | polyclonal | IgG |
| Tenascin C | 1:200 | 1:1,000 | mouse | monoclonal (EB2) | IgG1 |
| TSLP | 1:400 | 1:1,000 | rabbit | polyclonal | IgG |
| Uteroglobin |  | 1:1,000 | rabbit | polyclonal | IgG |
| ZO-1 | 1:200 | 1:1,000 | rabbit | polyclonal | IgG |
| Anti-rabbit, HRP-conjugated |  | 1:1,000 | goat | polyclonal | IgG |
| Anti-mouse, HRP-conjugated |  | 1:1,000 | goat | polyclonal | IgG |
| Alexa Fluor® 488 | 1:400 |  | goat (anti-rabbit or anti-mouse) | polyclonal | IgG |
| Alexa Fluor® 594 | 1:400 |  | goat (anti-rabbit or anti-mouse) | polyclonal | IgG |

**Table S2.** Primer sequences for qRT-PCR

Gene	Sequence (5’ — 3’)

ADAM33	ff 5’ - TCTTTCggATggAgCAgCTg

rv 5’ - gACgCTgTTTggTgTggTTC

α-SMA	ff 5’ - TgggCTCTgTAAggCCggCT

rv 5’ -CACCCCCTgATgTCTgggACg

GAPDH	ff 5’ - CTCTCTgCTCCTCCTgTTCgAC

rv 5’ - TgAGCgATgTggCTCggCT

FLG	ff 5’ - AAggAACTTCTggAAAAggAATTTC

rv 5’ - TTgTggTCTATATCCAAgTgATCCAT

FN	ff 5’ - ggTgACACTTATgAgCgTCCTAAAA

rv 5’ - AACATgTAACCACCAgTCTCATgTg

LIF	ff 5’ - ACAgAgCCTTTgCgTgAAAC

rv 5’ - TggTCCACACCAgCAgATAA

MUC5AC	ff 5’ - CCTTCgACggACAgAgCTAC

rv 5’ - TCTTgATggCCTTggAgCAg

PAR-2	ff 5’ - TCATTgTCACTgTCCTggCC

rv 5’ - AAgggTAgAgAggCAgAggg

SSCA-1	ff 5’ - ggAgCCAAAgACAACACTgC

rv 5’ - gCTTgTTggCgATCTTCAgC

Tenascin C	ff 5’ - TCAAAgACgTgCCAggAgAC

rv 5’ - CTgTCTgggAAACACgTCgA

TSLP	ff 5’ - CCCAggCTATTCggAAACTCAg

rv 5’ - CgCCACAATCCTTgTAATTgTg

Uteroglobin	ff 5’ - CCCCTCCTCCACCATgAAAC

rv 5’ - AAAgTTCCATggCAgCCTCA

ZO-1	ff 5’ - TCCTgCTTgACCTCCCTAAA

rv 5’ - ACAACACggAACACCTCTCC

TBX21	ff 5’ - TTgAggTgAACgACggAgAg

rv 5’ - CCAAggAATTgACAgTTgggT

GATA3	ff 5’ - gAACCggCCCCTCATTAAg

rv 5’ - ATTTTTCggTTTCTggTCTggAT

RORC	ff 5’ - CAATggAAgTggTgCTggTTAg

rv 5’ - gggAgTgggAgAAgTCAAAgAT

AHR	ff 5’ - CAAATCCTTCCAAgCggCATA

rv 5’ - CAAATCCTTCCAAgCggCATA

YHWAZ	ff 5’ - AgACggAAggTgCTgAgAAA

rv 5’ - gAAgCATTggggATCAAgAA

mTBX21	ff 5’ - TCAACCAgCACCAgACAgAgATg

rv 5’ - CACCAAgACCACATCCACAAACA

mGATA3	ff 5’ - AgAACCggCCCCTTATgAA

rv 5’ - AgTTCgCgCAggATgTCC

mRORC	ff 5’ - CCgCTgAgAgggCTTCAC

rv 5’ - TgCAggAgTAggCCACATTACA

mIL-22	ff 5’ - ggTgACgACCAgAACATCCAgA

rv 5’ - AgAgACATAAACAgCAggTCCAgT

mHPRT	ff 5’ - AggCCAgACTTTgTTggATTTgAA

rv 5’ - CAACTTgCgCTCATCTTAggCTTT

mYHWAZ	ff 5’ - AAgACAgCACgCTAATAATgC	rv 5’ - TTggAAggCCggTTAATTTTC

**Table S3. Highly differentially expressed proteins in healthy and atopic ECM as derived from semi-quantitative proteomics analysis**

| **Proteins and their function more abundant in healthy ECM** | | |
| --- | --- | --- |
| **TENA** | Tenascin | ECM Protein |
| **SODE** | Extracellular superoxide dismutase | Converts superoxide radicals into hydrogen peroxide and oxygen |
| **ROA2** | Heterogeneous nulear ribonucleoprotein A2/B1 | Transcription and processing of pre-mRNA |
| **NEST** | Nestin | Promotes disassembly of phosphorylated vimentin intermediate filaments (IF) during mitosis |
| **LUM** | Lumican | Organizes collagen fibril and ECM |
| **PGBM** | Basement-membrane specific heparan-sulfate proteoglycan core protein | Attachment substrate for cells playing an essential role in vascularization |
| **NID1** | Nidogen 1 | Organizes ECM and basement membrane |
| **G8JLB6** | Heterogeneous nuclear ribonucleoprotein H | RNA binding |
| **FINC** | Fibronectin | Involved in cell adhesion, cell motility, opsonization, wound healing, and maintenance |

| **Proteins and their function more abundant in atopic ECM** | | |
| --- | --- | --- |
| **SCOT1** | Succinyl-CoA:3-ketoacid coenzyme A transferase 1, mitochondrial | Key enzyme for ketone body catabolism |
| **DDR-2** | Discoidin domain-containing receptor 2 | Regulates remodeling of ECM |
| **ODO1** | 2-oxoglutarate dehydrogenase, mitochondrial | Mediates the decarboxylation of alpha-ketoglutarate |
| **THIK** | 3-ketoacyl-CoA thiolase, peroxisomal | Involved in thiolytic cleavage of straight chain 3-oxoacyl-CoAs |
| **MYOF** | Myoferlin | Calcium/phospholipid-binding protein |
| **VPP1** | V-type proton ATPase 116 kDa subunit a1 | Involved in ion membrane transport |
| **MFGM** | Lactadherin | Promotes VEGF-dependent neo-vascularization |
| **PYGB** | Glycogen phosphorylase, brain form | Regulates glycogen mobilization |
| **STAT1** | Signal transducer and activator of transcription 1-alpha/beta | Mediates cellular responses to interferons (IFNs), KITLG/SCF, other cytokines as well as growth factors |
| **TNPO1** | Transportin 1 | Nuclear transport receptor |
| **G6PD** | Glucose-6-phosphate 1-dehydrogenase | Provides reducing power (NADPH) and pentose phosphates for fatty acid and nucleic acid synthesis |
| **HTRA1** | Serine protease HTRA | Protease targeting ECM proteins |

**Table S4. Results of gene set enrichment analysis.** DB, name of the pathway collection; Contrast, the comparison for which the gene set enrichment was tested; ID, the identifier of the gene set in the MSigDB database; Title, name of the pathway; N1, number of genes in the pathway which were expressed in the analysed data set; AUC, effect size (area under curve); p.value, p-value from the CERNO test; FDR, p-value corrected for multiple testing using the Benjamini-Hochberg correction.

| DB | Contrast | ID | Title | N1 | AUC | P.Value | FDR |
| --- | --- | --- | --- | --- | --- | --- | --- |
| KEGG | CD44 vs PBS | M18009 | KEGG_CIRCADIAN_RHYTHM_MAMMAL | 13 | 0.8736 | 1.91e-11 | 3.553e-09 |
| ,, | ,, | M13088 | KEGG_PPAR_SIGNALING_PATHWAY | 52 | 0.7048 | 3.465e-07 | 3.222e-05 |
| ,, | ,, | M12868 | KEGG_PATHWAYS_IN_CANCER | 291 | 0.5529 | 7.895e-06 | 0.0004895 |
| ,, | ,, | M3985 | KEGG_CITRATE_CYCLE_TCA_CYCLE | 28 | 0.7213 | 9.509e-05 | 0.004422 |
| ,, | ,, | M16004 | KEGG_ANTIGEN_PROCESSING_AND_PRESENTATION | 52 | 0.6 | 0.0001947 | 0.007243 |
| ,, | ,, | M11521 | KEGG_GLYCOLYSIS_GLUCONEOGENESIS | 46 | 0.6521 | 0.0004594 | 0.01068 |
| ,, | ,, | M19540 | KEGG_OXIDATIVE_PHOSPHORYLATION | 115 | 0.6248 | 0.0003889 | 0.01068 |
| ,, | ,, | M7934 | KEGG_PYRUVATE_METABOLISM | 32 | 0.6724 | 0.0004509 | 0.01068 |
| ,, | ,, | M17673 | KEGG_CARDIAC_MUSCLE_CONTRACTION | 60 | 0.6665 | 0.0009717 | 0.02008 |
| ,, | ,, | M13950 | KEGG_ASTHMA | 19 | 0.6805 | 0.001287 | 0.02302 |
| ,, | ,, | M1835 | KEGG_GLIOMA | 61 | 0.5631 | 0.001362 | 0.02302 |
| ,, | ,, | M16473 | KEGG_ALDOSTERONE_REGULATED_SODIUM_REABSORPTION | 36 | 0.579 | 0.003745 | 0.04976 |
| ,, | ,, | M16024 | KEGG_ALZHEIMERS_DISEASE | 150 | 0.5916 | 0.00327 | 0.04976 |
| ,, | ,, | M980 | KEGG_TRYPTOPHAN_METABOLISM | 28 | 0.6862 | 0.003674 | 0.04976 |
| ,, | SD4 vs PBS | M18009 | KEGG_CIRCADIAN_RHYTHM_MAMMAL | 13 | 0.8696 | 2.398e-15 | 4.46e-13 |
| ,, | TSP1 vs PBS | M18009 | KEGG_CIRCADIAN_RHYTHM_MAMMAL | 13 | 0.8655 | 1.507e-09 | 2.803e-07 |
| ,, | ,, | M13191 | KEGG_PROSTATE_CANCER | 84 | 0.599 | 0.0002103 | 0.01956 |
| REACTOME | CD44 vs PBS | M938 | REACTOME_CIRCADIAN_CLOCK | 66 | 0.5889 | 4.66e-07 | 0.0007475 |
| ,, | ,, | M27451 | REACTOME_METABOLISM_OF_LIPIDS | 604 | 0.5563 | 1.095e-06 | 0.0008781 |
| ,, | ,, | M8276 | REACTOME_NUCLEAR_RECEPTOR_TRANSCRIPTION_PATHWAY | 43 | 0.6405 | 3.088e-06 | 0.001651 |
| ,, | ,, | M27332 | REACTOME_CALCITONIN_LIKE_LIGAND_RECEPTORS | 8 | 0.767 | 2.896e-05 | 0.01066 |
| ,, | ,, | M490 | REACTOME_PYRUVATE_METABOLISM_AND_CITRIC_ACID_TCA_CYCLE | 49 | 0.6794 | 3.989e-05 | 0.01066 |
| ,, | ,, | M516 | REACTOME_THE_CITRIC_ACID_TCA_CYCLE_AND_RESPIRATORY_ELECTRON_TRANSPORT | 163 | 0.6186 | 3.841e-05 | 0.01066 |
| ,, | ,, | M27854 | REACTOME_FATTY_ACID_METABOLISM | 145 | 0.6047 | 5.874e-05 | 0.01346 |
| ,, | ,, | M729 | REACTOME_FATTY_ACYL_COA_BIOSYNTHESIS | 30 | 0.6688 | 7.171e-05 | 0.01438 |
| ,, | ,, | M17157 | REACTOME_PYRUVATE_METABOLISM | 25 | 0.7393 | 0.0001065 | 0.01897 |
| ,, | ,, | M27946 | REACTOME_FOXO_MEDIATED_TRANSCRIPTION_OF_CELL_CYCLE_GENES | 15 | 0.6651 | 0.0001638 | 0.02627 |
| ,, | ,, | M27316 | REACTOME_REGULATION_OF_LIPID_METABOLISM_BY_PPARALPHA | 107 | 0.5457 | 0.0002441 | 0.03263 |
| ,, | ,, | M38997 | REACTOME_SIGNALING_BY_MAPK_MUTANTS | 7 | 0.8201 | 0.0002421 | 0.03263 |
| ,, | ,, | M797 | REACTOME_CLASS_B_2_SECRETIN_FAMILY_RECEPTORS | 54 | 0.6108 | 0.0003017 | 0.03623 |
| ,, | ,, | M746 | REACTOME_SIGNALING_BY_GPCR | 425 | 0.5303 | 0.0003162 | 0.03623 |
| ,, | ,, | M27609 | REACTOME_INTERLEUKIN_4_AND_INTERLEUKIN_13_SIGNALING | 92 | 0.5448 | 4e-04 | 0.04277 |
| ,, | ,, | M26943 | REACTOME_BMAL1_CLOCK_NPAS2_ACTIVATES_CIRCADIAN_GENE_EXPRESSION | 24 | 0.6283 | 0.0004474 | 0.04485 |
| ,, | SD4 vs PBS | M938 | REACTOME_CIRCADIAN_CLOCK | 66 | 0.5897 | 3.634e-08 | 5.829e-05 |
| ,, | ,, | M738 | REACTOME_PHASE_I_FUNCTIONALIZATION_OF_COMPOUNDS | 65 | 0.6848 | 6.749e-07 | 0.0005413 |
| ,, | ,, | M26943 | REACTOME_BMAL1_CLOCK_NPAS2_ACTIVATES_CIRCADIAN_GENE_EXPRESSION | 24 | 0.6399 | 6.561e-06 | 0.003005 |
| ,, | ,, | M5650 | REACTOME_CYTOCHROME_P450_ARRANGED_BY_SUBSTRATE_TYPE | 32 | 0.7202 | 7.494e-06 | 0.003005 |
| ,, | ,, | M8276 | REACTOME_NUCLEAR_RECEPTOR_TRANSCRIPTION_PATHWAY | 43 | 0.5774 | 0.0001136 | 0.03645 |

**Figures**
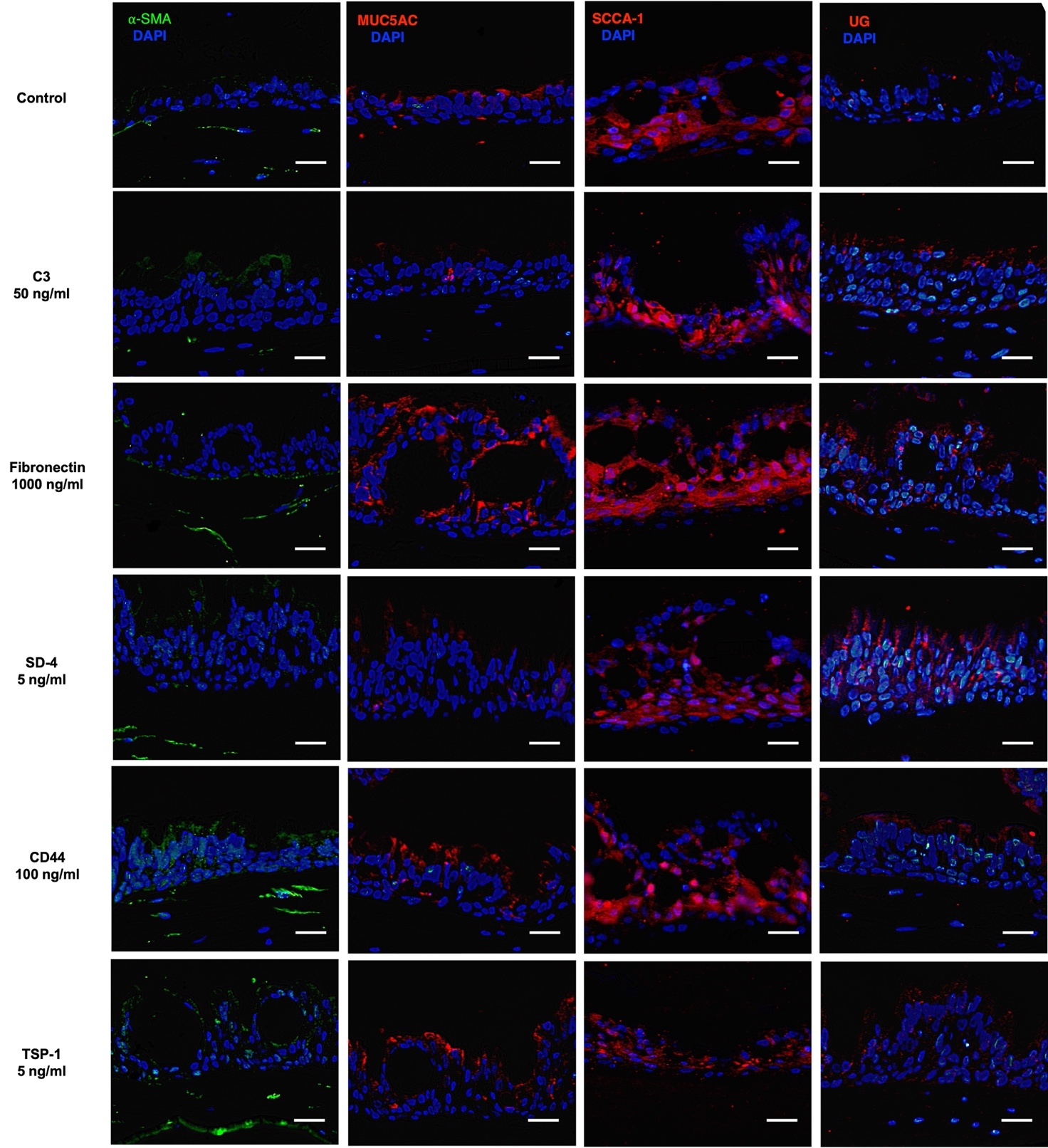

**Figure S1. Immunofluorescence staining:** Bronchial epithelial equivalents were either treated with 50 ng/ml complement factor C3 (C3), 1000 ng/ml fibronectin, 5 ng/ml syndecan-4 (SD-4), 100 ng/ml soluble cluster of differentiation 44 (CD44), or 5 ng/ml thrombospondin-1 (TSP-1) for 6 days. Representative pictures of IF-staining for, alpha smooth muscle actin (𝛼-SMA), MUC5AC, SCCA-1, and uteroglobin (UG) are shown. Exposure times: blue channel 1/15s, red channel 1/3s, green channel 1/3s. Scale bar = 25µm.

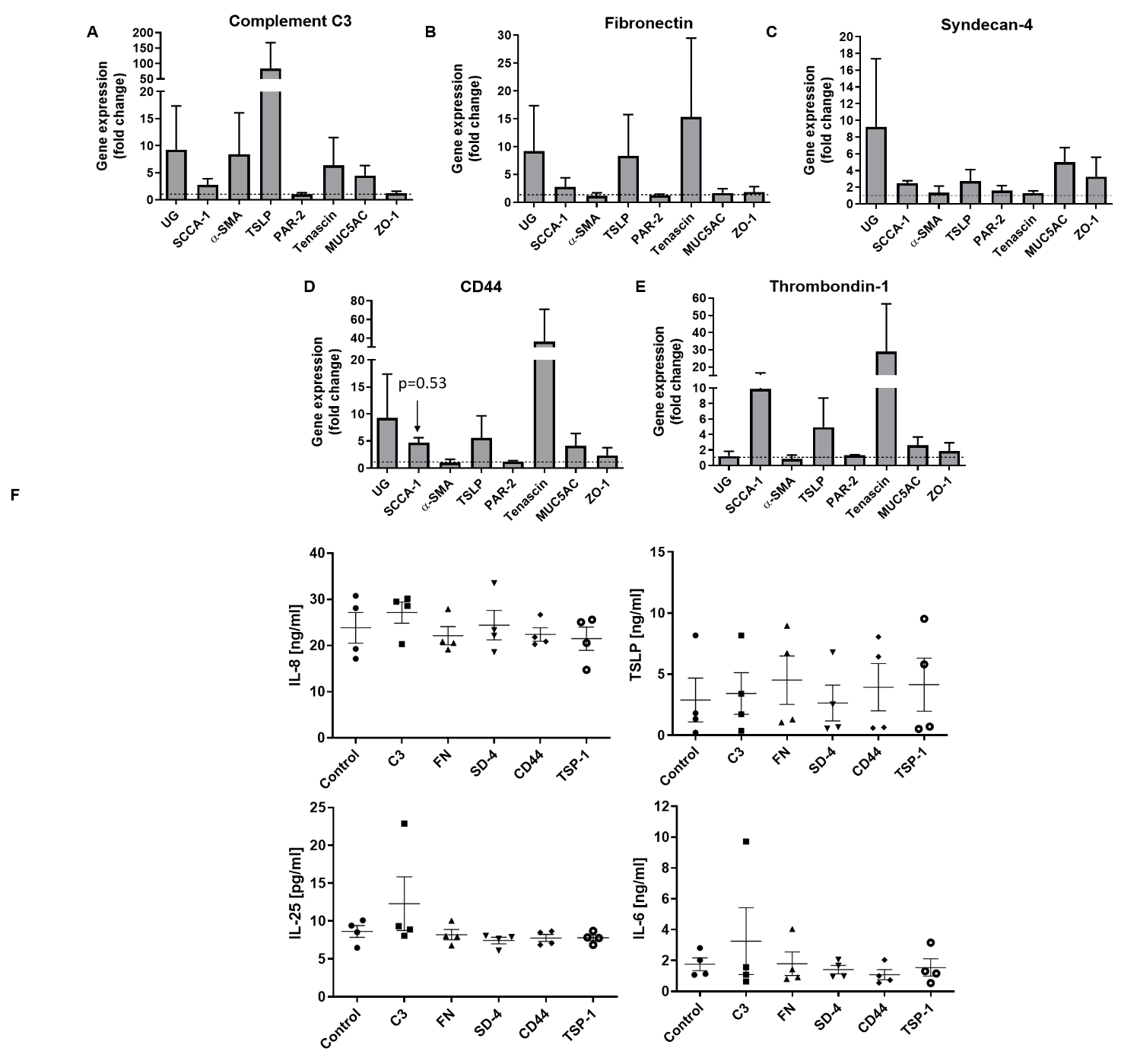

**Figure S2. Gene expression and cytokine release after treating bronchial epithelial models with A)** 50 ng/ml C3, **B)** 1μg/ml FN, **C)** 5 ng/ml SD-4, **D** )100 ng/ml CD-44, or **E)** 5 ng/ml TSP-1 over 6 days as determined by qRT-PCR. mRNA-expression of uteroglobin (*UG*), *SCCA-1*, alpha smooth muscle actin (*𝛼-SMA*), *TSLP*, *PAR-2*, tenascin C, *MUC5AC*, and *ZO-1* in bronchial epithelial equivalents after a 6-day treatment with. GAPDH served as housekeeping gene. Mean ± SEM of fold change. n=3. **F** Secretion of IL-6, IL-8, TSLP, and IL-25 into culture media of bronchial epithelial models after 6-days treatment with each compound as determined by sandwich ELISA. n=4 independent donors.

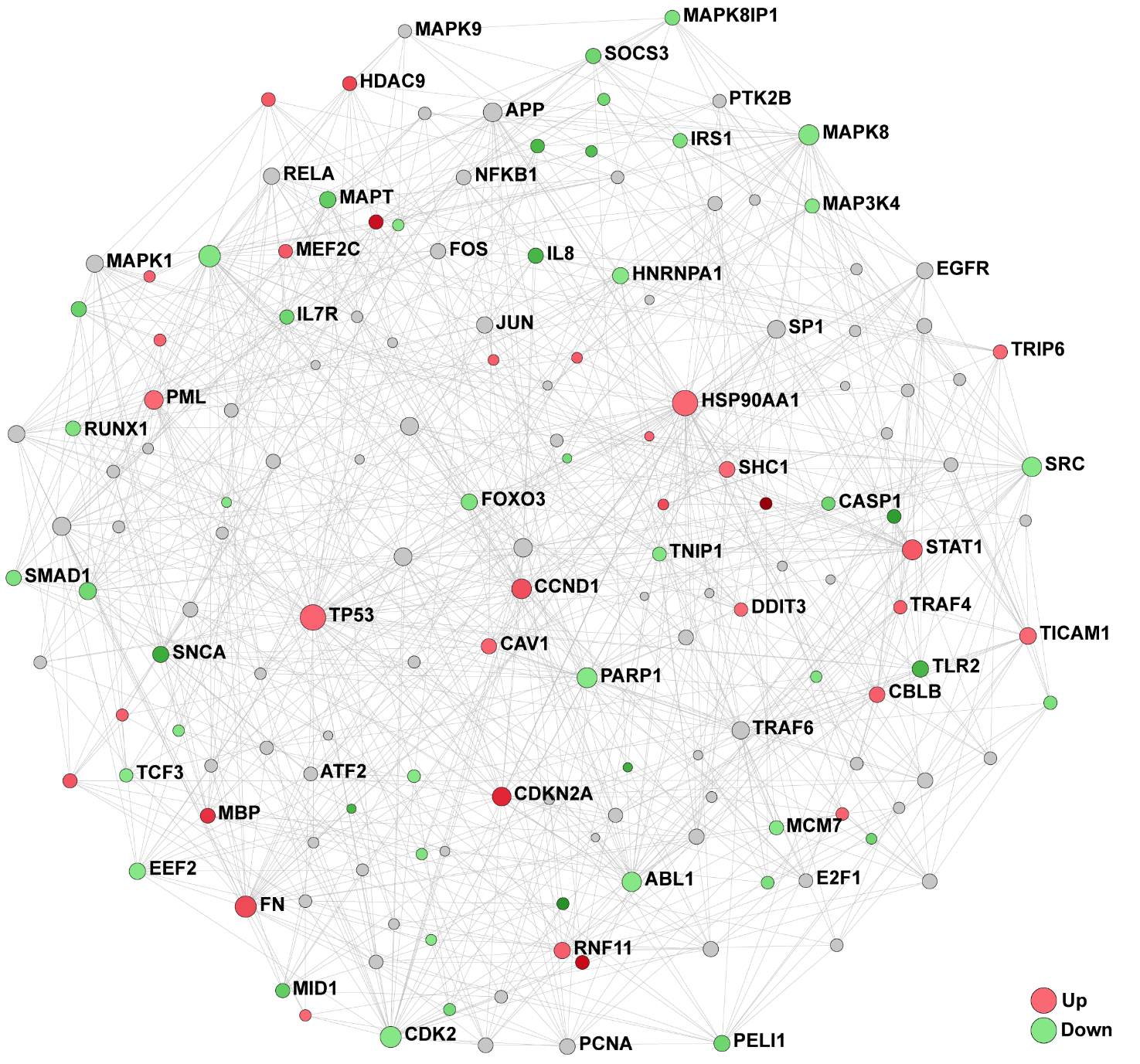

**Figure S3. Transcriptomic analysis of healthy and AD fibroblasts:** Regulation of TLR4 signaling network.

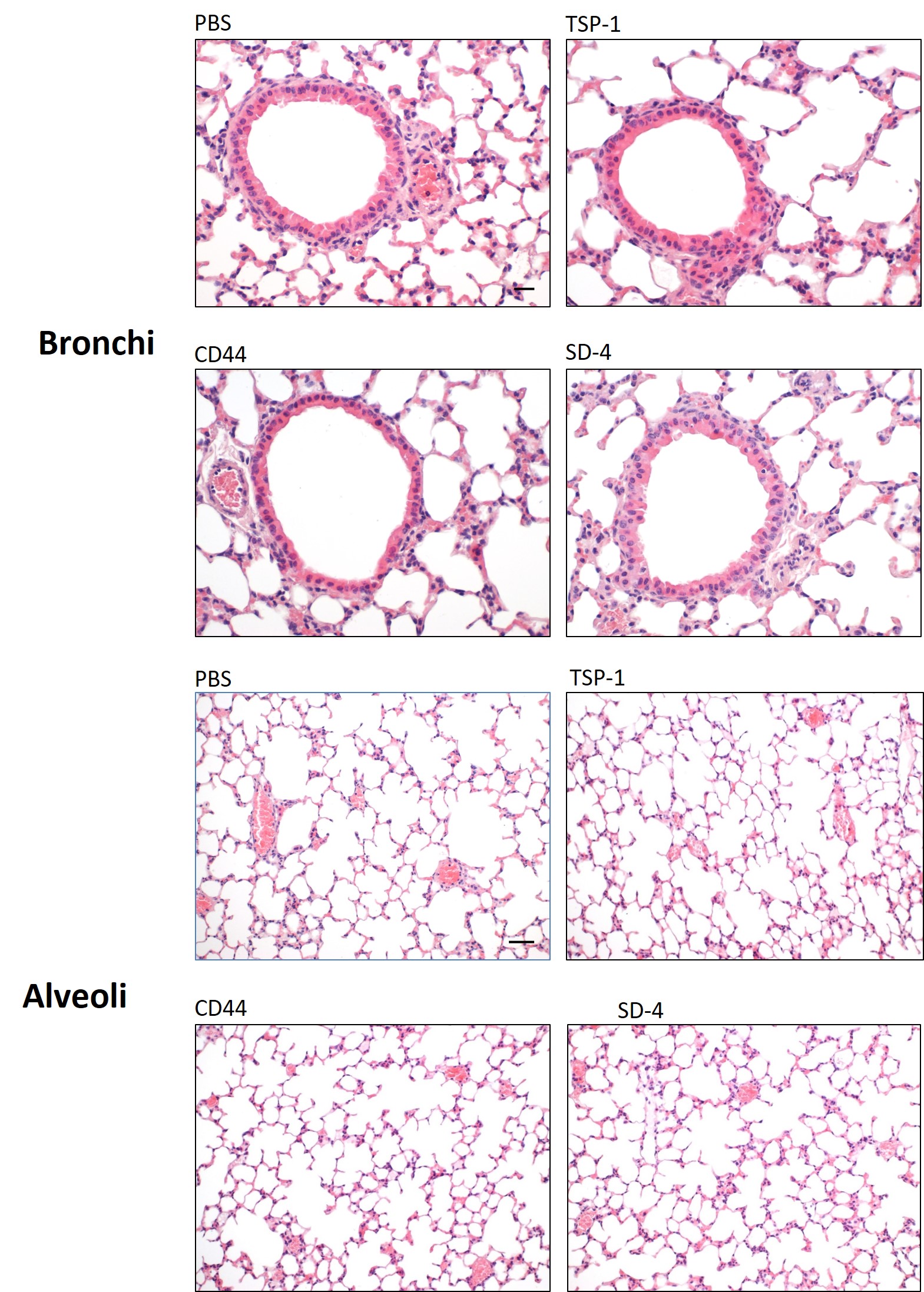

**Figure S4. Histology of mice lungs:** Representative H&E-staining of bronchi and alveoli of control (PBS), TSP-1, CD44, and SD-4 treated mice after 7 days. Scale bars: Bronchi = 20 µm, alveoli = 50 µm

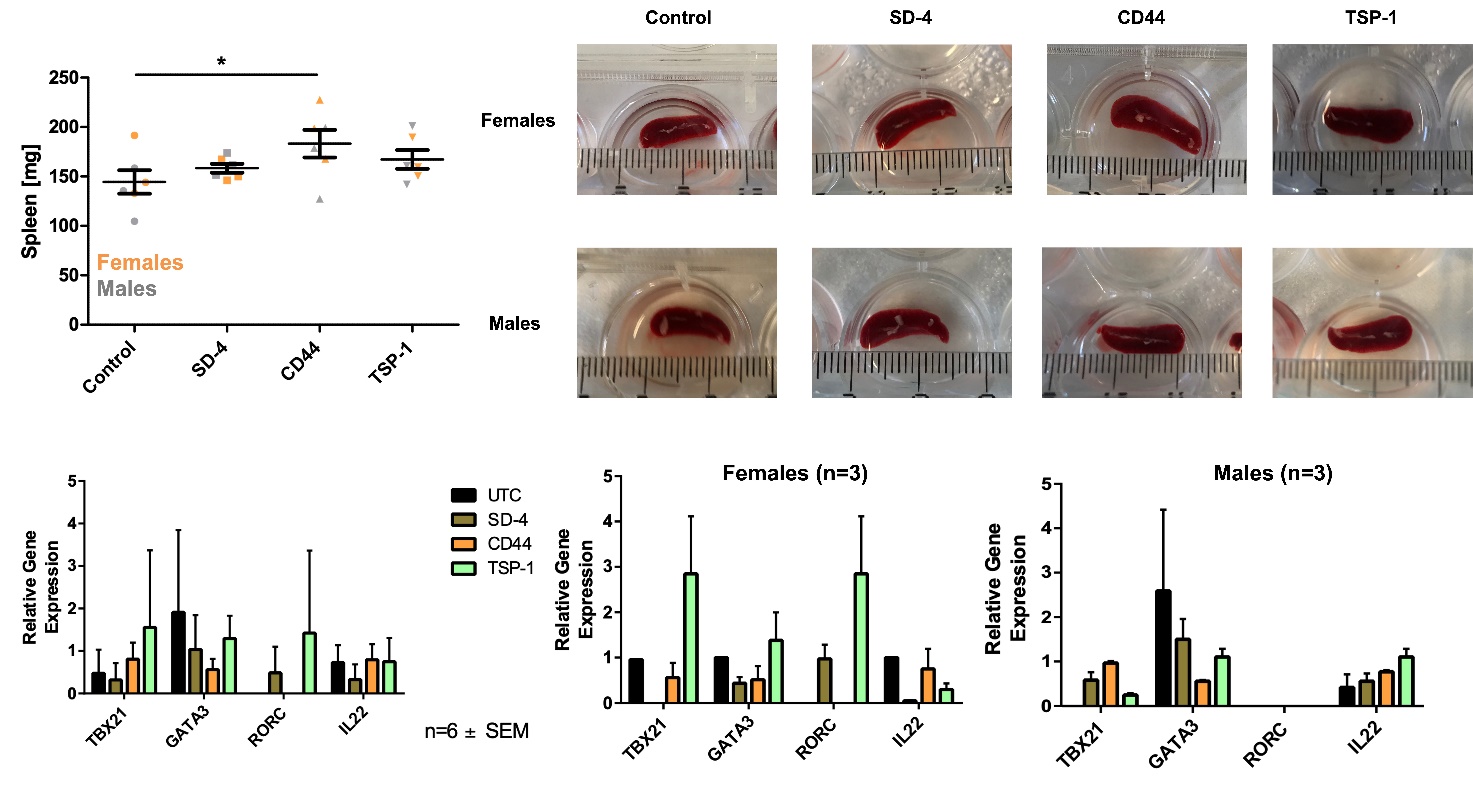
**Figure S5. Spleen weight and size after treatment and gene expression in murine CD4^+^ T cells.** **Upper panel**: After 7-day treatment of mice with SD-4, CD44, or TSP-1 the spleens were removed, weighed and measured. A dot plot shows the average weight of spleen from female (orange) and male (grey) mice. Mean ± SEM. n=6. *p≤0.05 as determined by a one-Way-Anova with Dunnett’s Multiple Comparison Post-Test.

**Lower Panel:** CD4^+^ T cells were isolated from mouse spleens after 7-day treatment with SD-4, CD44, or TSP-1 and mRNA expression of TBX21 (Th1), GATA3 (Th2), RORC (Th17), and IL-22 (Th22) of both female and male mice was determined via qRT-PCR. GAPDH and HPRT served as housekeeping genes. Mean ± SEM. n=6. Left: pooled data, right: sex-disaggregated data.

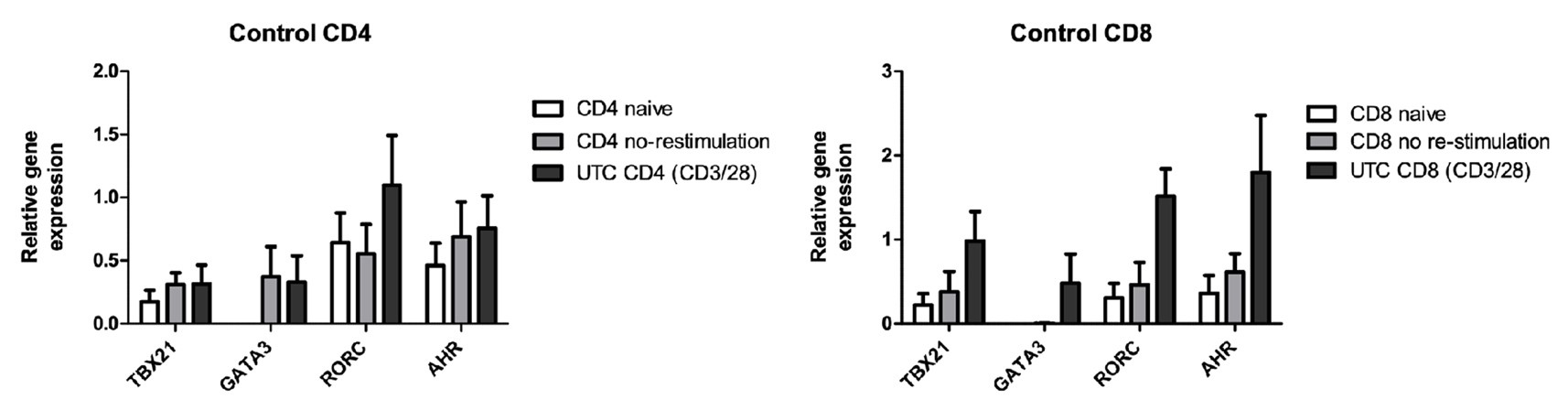

**Figure S6. Gene expression of CD4^+^ and CD8^+^ T cells:** After isolation of naive CD4^+^ and naive CD8+ T cells from PBMCs T cells were stimulated with CD3/ CD28 for two days and re-stimulated with PMA/ionomycin at day 12. The normalized expression of *TBX21*, *GATA3*, *RORC*, and *AHR* as determined by qRT-PCR is shown. GAPDH served as housekeeping gene. Mean ± SEM. n=4 independent donors.

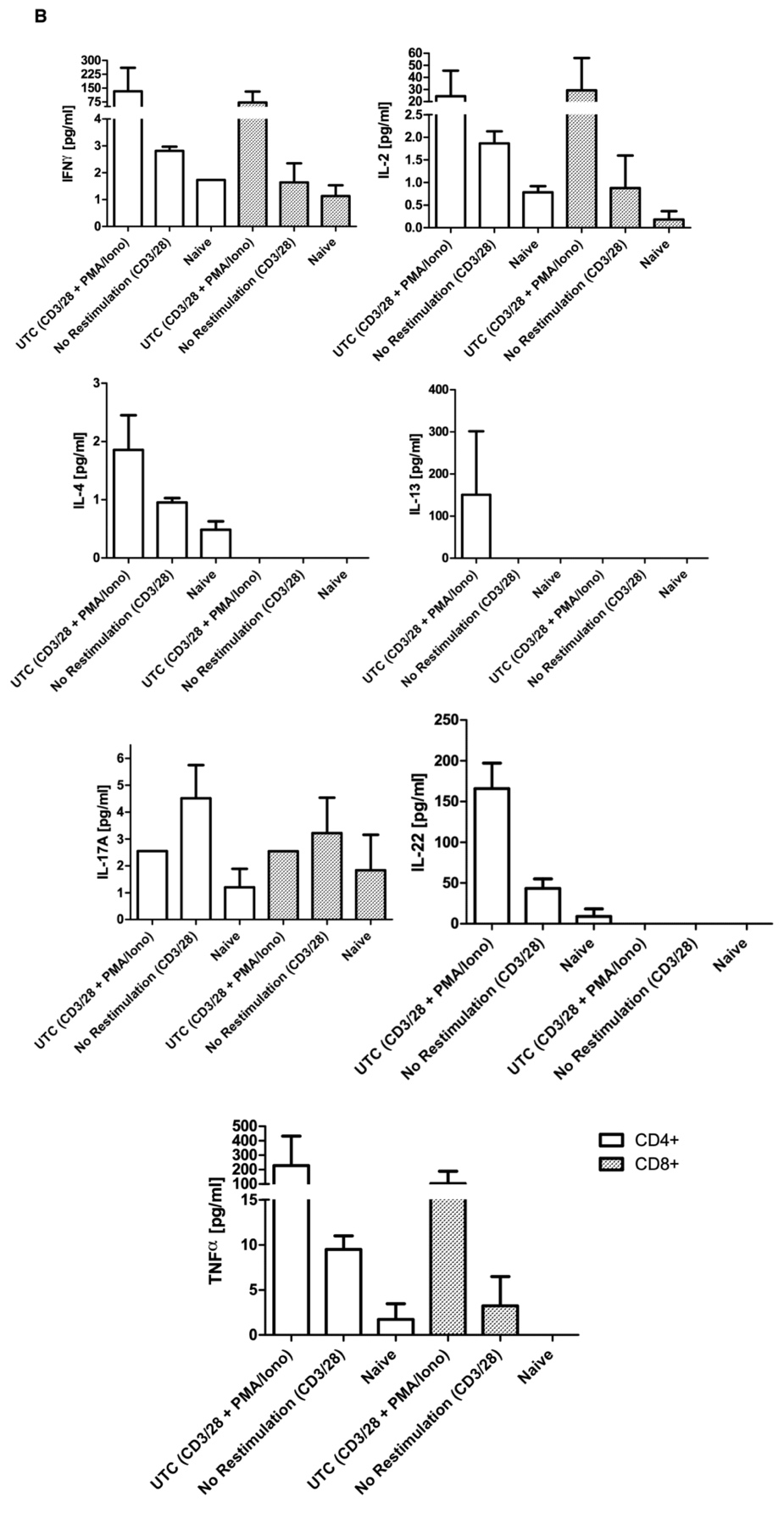

**Figure S7.** Untreated, not re-stimulated, and naive controls for CD4^+^ and CD8^+^ T cells. Mean ± SEM. n=2 independent donors.

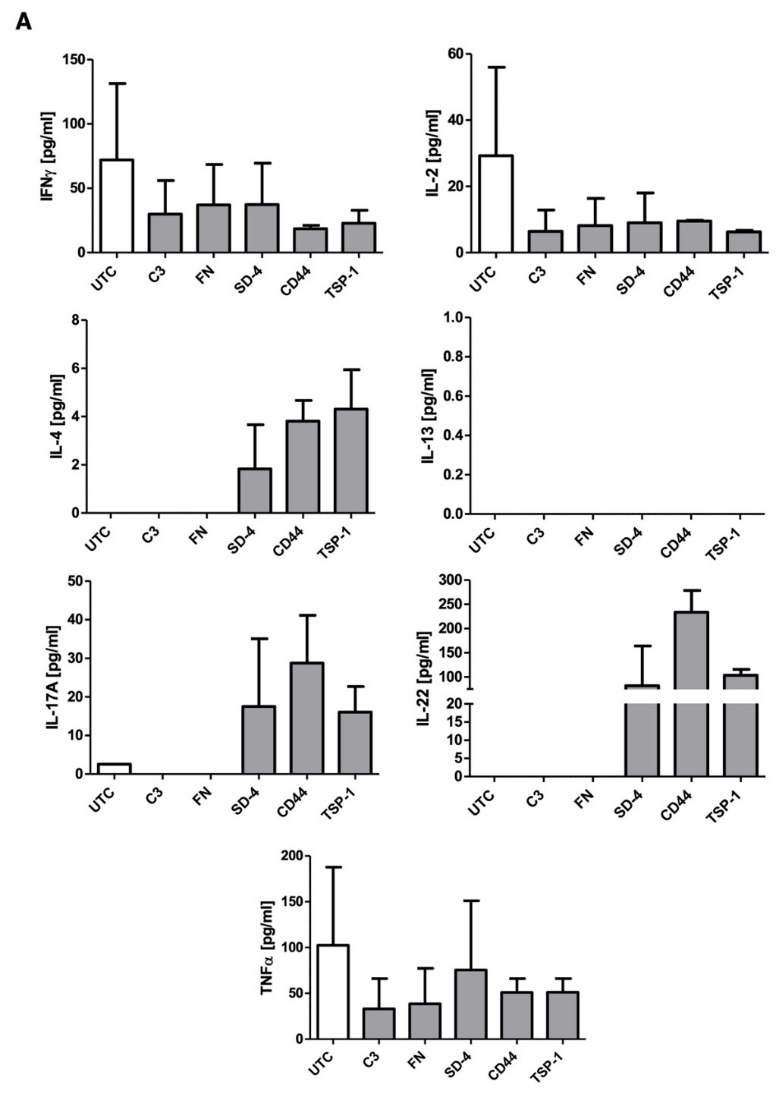

**Figure S8. Multiplex ELISA assay of human CD8^+^ T cells** after incubation with C3, fibronectin (FN), syndecan-4 (SD-4), CD44, and thrombospondin-1 (TSP-1) over 10 days.

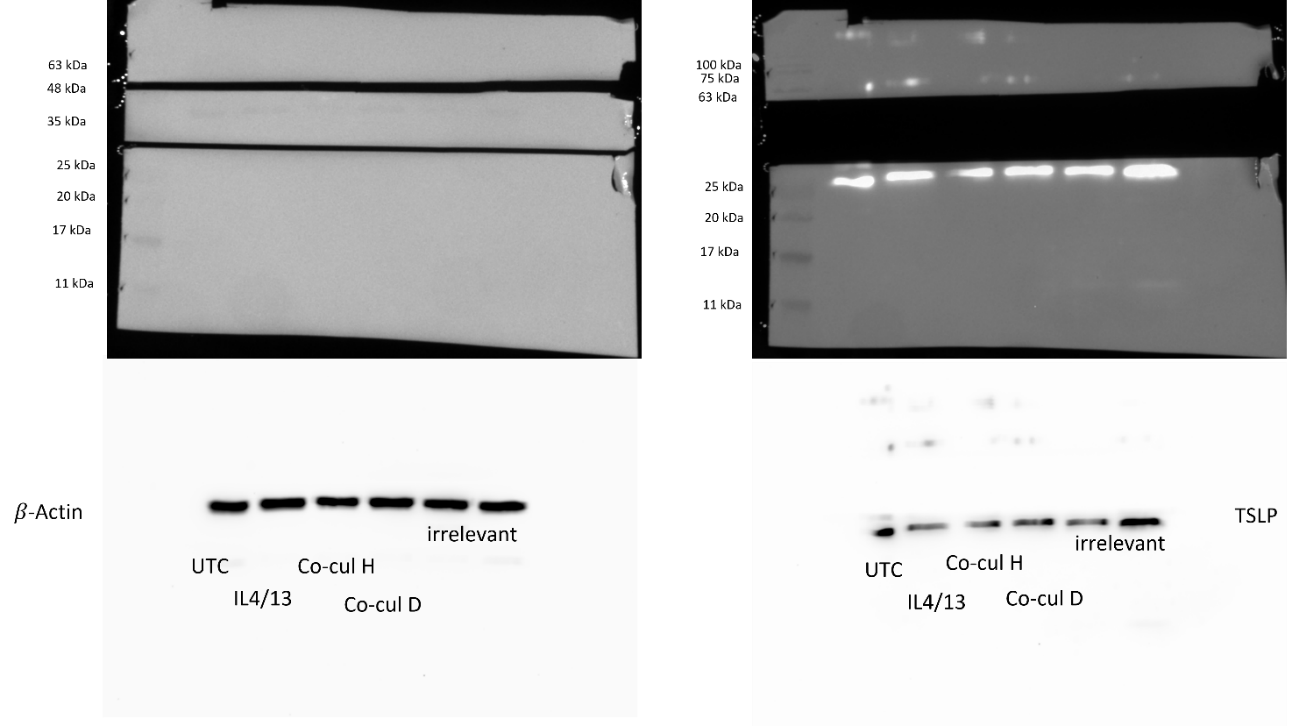

**Figure S9. Exemplary western blot membrane:** Membranes were cut twice horizontally below 35 kDa and below 63 kDa. Exemplary blots of co-culture experiments are shown for TSLP (28 kDa) and 𝛽-Actin (42 kDa).

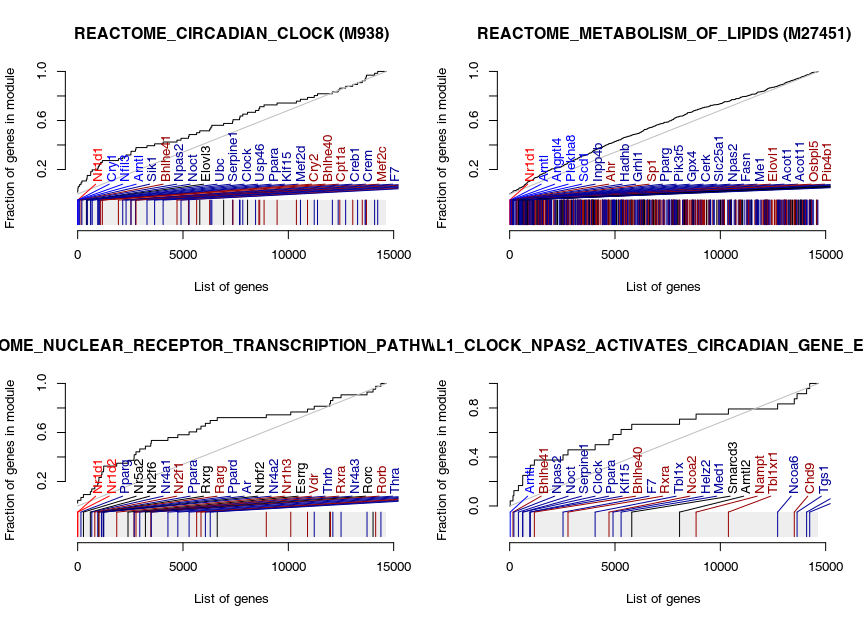

**Figure S10.** Evidence plots for selected gene sets in the CD44 contrast. The curve is a receiver operator characteristic (ROC) curve of the enrichments. Grey rug corresponds to a list of genes ordered by their p-values. Position of genes belonging to the given gene sets is indicated; colors correspond to the log2 fold change. Red indicates log2 fold change greater than zero; blue indicates log2 fold change smaller than zero. Bright colors indicate genes significant at FDR < 0.05. Gene symbols are shown for the first 30 genes.

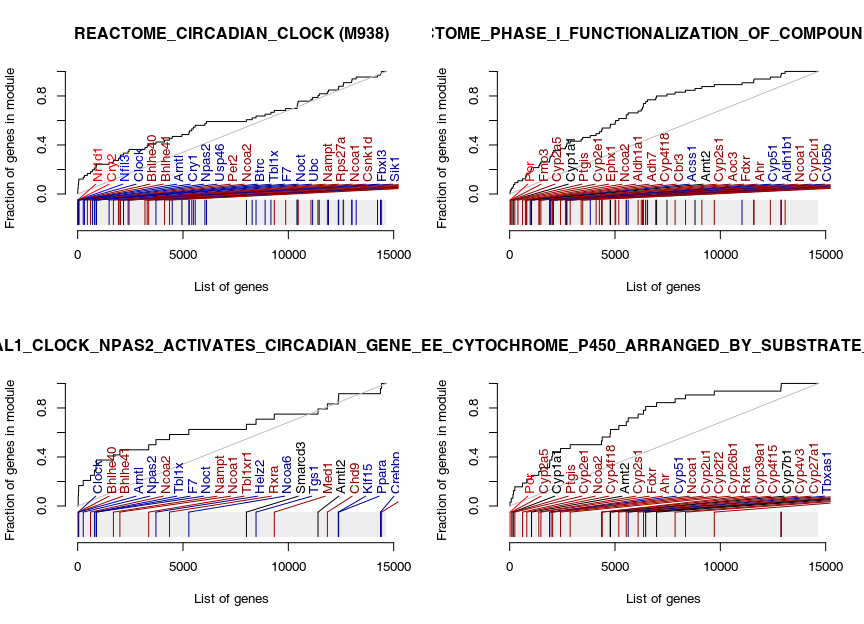

**Figure S11.** Evidence plots for selected gene sets in the SD4 contrast. The curve is a receiver operator characteristic (ROC) curve of the enrichments. Grey rug corresponds to a list of genes ordered by their p-values. Position of genes belonging to the given gene sets is indicated; colors correspond to the log2 fold change. Red indicates log2 fold change greater than zero; blue indicates log2 fold change smaller than zero. Bright colors indicate genes significant at FDR < 0.05. Gene symbols are shown for the first 30 genes.

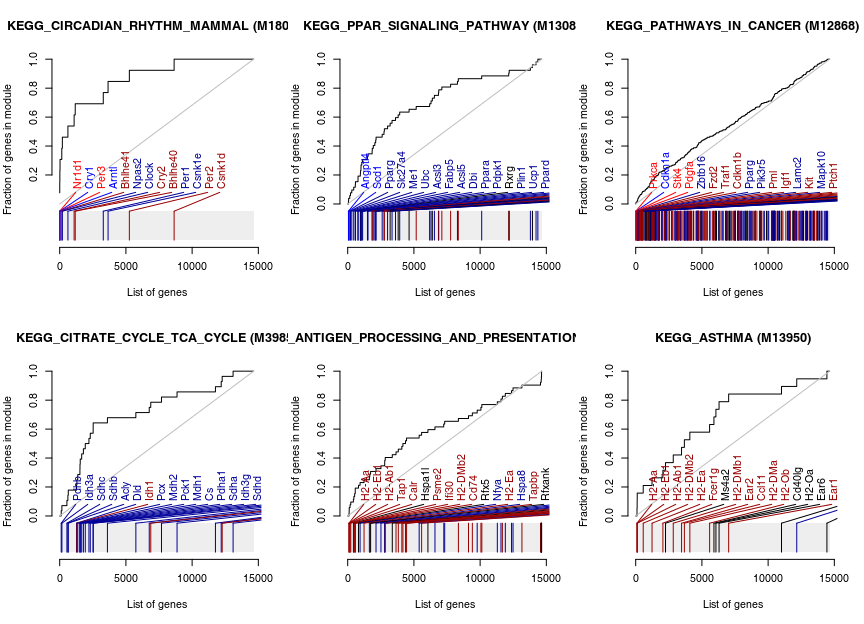

**Figure S12.** Evidence plots for selected gene sets in the CD44 (db msigdb_kegg) contrast. The curve is a receiver operator characteristic (ROC) curve of the enrichments. Grey rug corresponds to a list of genes ordered by their p-values. Position of genes belonging to the given gene sets is indicated; colors correspond to the log2 fold change. Red indicates log2 fold change greater than zero; blue indicates log2 fold change smaller than zero. Bright colors indicate genes significant at FDR < 0.05. Gene symbols are shown for the first 30 genes.

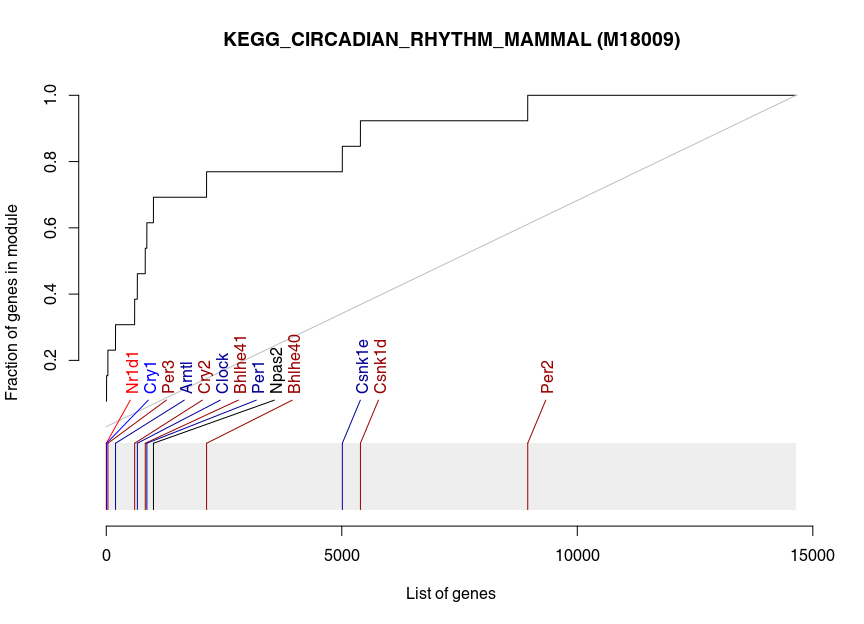

**Figure S13.** Evidence plots for selected gene sets in the TSP1(db msigdb_kegg) contrast. The curve is a receiver operator characteristic (ROC) curve of the enrichments. Grey rug corresponds to a list of genes ordered by their p-values. Position of genes belonging to the given gene sets is indicated; colors correspond to the log2 fold change. Red indicates log2 fold change greater than zero; blue indicates log2 fold change smaller than zero. Bright colors indicate genes significant at FDR < 0.05. Gene symbols are shown for the first 30 genes.

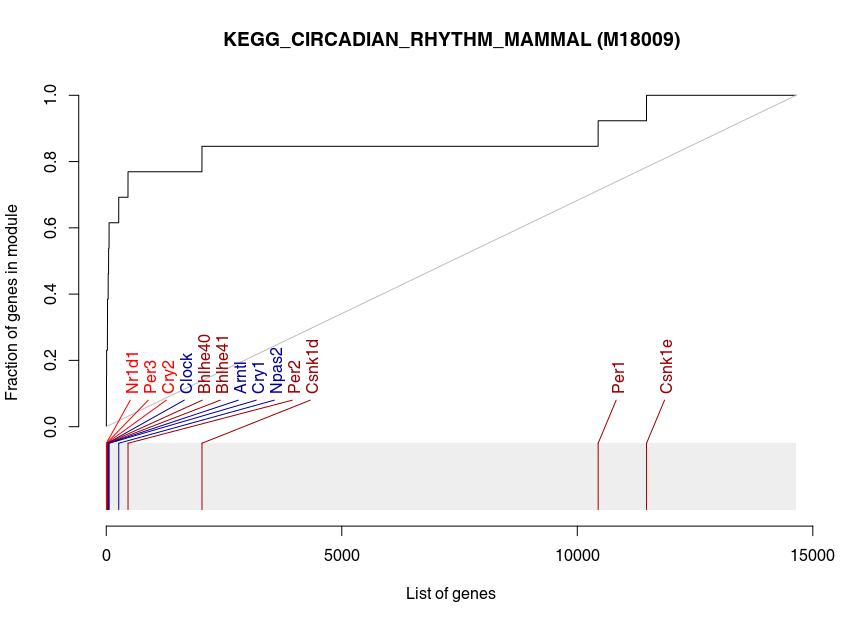

**Figure S14.** Evidence plots for selected gene sets in the SD4 (db msigdb_kegg) contrast. The curve is a receiver operator characteristic (ROC) curve of the enrichments. Grey rug corresponds to a list of genes ordered by their p-values. Position of genes belonging to the given gene sets is indicated; colors correspond to the log2 fold change. Red indicates log2 fold change greater than zero; blue indicates log2 fold change smaller than zero. Bright colors indicate genes significant at FDR < 0.05. Gene symbols are shown for the first 30 genes.

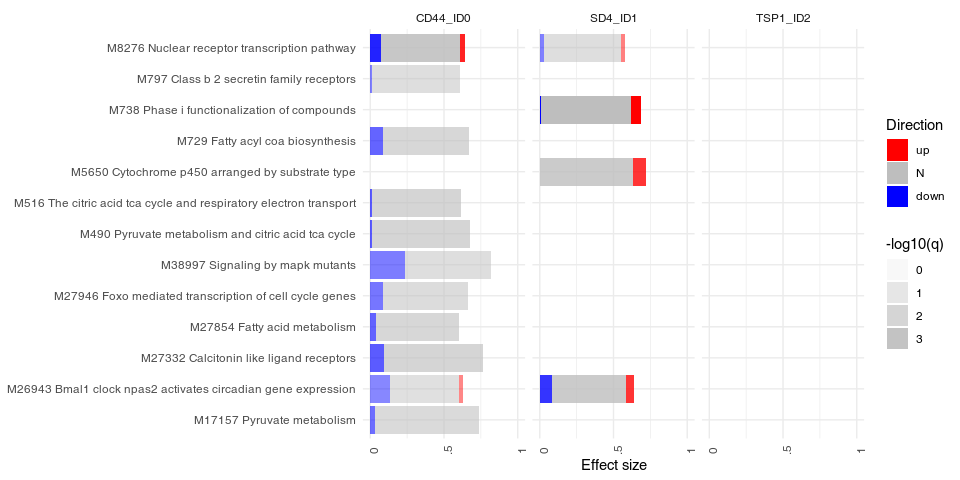

**Figure S15.** Overview of the results of gene set enrichment analysis for the database REACTOME.

**References**

1.	Vávrová, K., et al., *Filaggrin deficiency leads to impaired lipid profile and altered acidification pathways in a 3D skin construct.* J Invest Dermatol, 2014. **134**(3): p. 746-53.

2.	Hönzke, S., et al., *Influence of Th2 Cytokines on the Cornified Envelope, Tight Junction Proteins, and ß-Defensins in Filaggrin-Deficient Skin Equivalents.* J Invest Dermatol, 2016. **136**(3): p. 631-9.

3.	Lu, H., et al., *Subcutaneous Angiotensin II Infusion using Osmotic Pumps Induces Aortic Aneurysms in Mice.* J Vis Exp, 2015(103).

4.	Bäumer, W., et al., *Cilomilast, tacrolimus and rapamycin modulate dendritic cell function in the elicitation phase of allergic contact dermatitis.* Br J Dermatol, 2005. **153**(1): p. 136-44.
